# Forecasting COVID-19 with Temporal Hierarchies and Ensemble Methods

**DOI:** 10.1101/2025.06.26.25330355

**Authors:** Li Shandross, Evan L. Ray, Benjamin W. Rogers, Nicholas G. Reich

## Abstract

Infectious disease forecasting efforts underwent rapid growth during the COVID-19 pandemic, providing guidance for pandemic response and about potential future trends. Yet despite their importance, short-term forecasting models often struggled to produce accurate real-time predictions of this complex and rapidly changing system. This gap in accuracy persisted into the pandemic and warrants the exploration and testing of new methods to glean fresh insights.

In this work, we examined the application of the temporal hierarchical forecasting (THieF) methodology to probabilistic forecasts of COVID-19 incident hospital admissions in the United States. THieF is an innovative forecasting technique that aggregates time-series data into a hierarchy made up of different temporal scales, produces forecasts at each level of the hierarchy, then reconciles those forecasts using optimized weighted forecast combination. Vhile THieF’s unique approach has shown substantial accuracy improvements in a diverse range of applications, such as operations management and emergency room admission predictions, this technique had not previously been applied to outbreak forecasting.

We generated candidate models formulated using the THieF methodology, which differed by their hierarchy schemes and data transformations, and ensembles of the THieF models, computed as a mean of predictive quantiles. The models were evaluated using weighted interval score (WIS) as a measure of forecast skill, and the top-performing subset was compared to a group of benchmark models. These models included simple ARIMA and seasonal ARIMA models, an ensemble of these ARIMA models, a naive baseline model, four operational incident hospitalization models from the U.S. COVID-19 Forecast Hub, and an equally-weighted quantile median of all models that submitted incident hospitalization forecasts to the Forecast Hub. The THieF models and THieF ensembles demonstrated improvements in WIS and MAE, as well as competitive prediction interval coverage, over many benchmark models for both the validation and testing phases. The best THieF model’s rank oscillated between second or third out of fourteen total models during the testing evaluation. These accuracy improvements suggest the THieF methodology may serve as a useful addition to the infectious disease forecasting toolkit.

## 1 Introduction

During the pandemic, predictions of COVID-19 dynamics informed public policy through balancing prevention and mitigation efforts with other societal and monetary costs while also guiding proper allocation of resources [22]. Infectious disease forecasting saw rapid growth due to its widespread usage during this time. However, many COVID-19 models struggled with creating accurate forecasts, particularly when predicting trend fluctuations, in which the slope or direction of the data deviate from the previously held pattern [8]. This gap clearly necessitates the exploration and testing of new methods.

Temporal hierarchical forecasting THieF) is one such method that uniquely combines hierarchical forecasting and forecast combination for time series data. This approach, developed by Athanasopoulos, et al. in 2017, consists of three main steps: aggregation of data at various temporal resolutions to create a hierarchy, generation of forecasts at each level of the hierarchy, and reconciliation of those forecasts. The final step performs weighted forecast combination to not only achieve coherence but also improved accuracy [1]. Coherence describes when hierarchical point forecasts predict the same trend, event, or outcome at every level such that the forecasts at lower levels add up to those at higher levels. Reconciliation describes the process of making forecasts coherent. Vhen forecasts from different levels of the hierarchy disagree about the future are not coherent), it is unclear which level s forecasts should be trusted and chosen to inform decision-making [14].

We propose that the THieF methodology may have something to offer to COVID-19 forecasting, and the field of infectious disease forecasting as a whole. First, THieF relies on a phenomenological model, a class of models that use only observed data to make predictions, which have shown good performance for short-term outbreak forecasting [13]. Second, to our knowledge, THieF has yet to be applied to outbreak forecasting. Hence, this methodology may offer insights into effective modeling techniques. Third, THieF has shown significant performance gains in other settings, which may translate to infectious disease forecasting [8, 1].

We ensemble the THieF models to take advantage of the performance benefits of forecast combination. Research has shown that aggregating predictions from multiple sources produce forecasts that are more accurate and consistent than individual models [3, 23, 11]. The field of infectious disease forecasting has also recently adopted the usage of multi-model ensembles to improve predictions of disease outbreaks, and a simple, equally-weighted median ensemble has been found to be one of the most accurate COVID-19 models for predicting deaths 16, 8.

In this work, we use models constructed with the THieF methodology and ensembles of the THieF models to retrospectively forecast for COVID-19 daily incident hospitalizations in the United States. These original THieF models, the THieF ensembles, a group of Autoregressive Integrated Moving Average (ARIMA) and seasonal ARIMA (SARIMA) models, and an equally-weighted mean ensemble of the ARIMA and SARIMA models make up a total of twenty-seven (27) candidate models evaluated during the validation phase. A most-accurate subset of up to ten (10) models are passed on to the testing phase where they are compared to six benchmark models: four operational models that submitted incident hospitalization predictions to the U.S. COVID-19 Forecast Hub (shortened to the “Forecast Hub” from here on), a naive baseline model, and an untrained quantile median ensemble of all models from the Forecast Hub that predicted incident hospitalizations. The Forecast Hub is a centralized repository that synthesizes, collects, archives, and evaluates COVID-19 forecasts [7]. More information on the selected benchmark models can be found in subsection [2.4].

Success is demonstrated by a few original THieF models and the THieF ensembles, which showed substantial accuracy gains over the baseline and three of the four operational models. Further, one of the THieF models also displayed modest improvements over a top-performing equally-weighted median ensemble when the number of incident hospitalizations was changing rapidly from week to week.

## 2 Methods

### 2.1 Surveillance data

We used confirmed hospital admissions (aggregated by state daily) reported by HHS Protect to train and evaluate our models [24]. Our choice of data source was motivated by its designation as the official ground truth data for COVID-19 incident hospitalizations by the Forecast Hub [7]. The choice to forecast incident hospitalizations was motivated by the target s more consistent and reliable values (due to reporting requirements for Medicare reimbursement) compared to other COVID-19 forecasting targets. For example, incident cases and incident deaths have suffered from massive under reporting and are thus less relevant public health metrics than incident hospitalizations [10, 9, 13]. Surveillance data, like the HHS Protect confirmed hospitalizations, sometimes requires revisions due to reporting anomalies. Hospitalizations might be entered on incorrect dates or added after COVID-19 was determined to be the cause of hospitalization. Updates to values usually occurred within the first week or so of posting. As a result, real-time forecast models might train on data that include incident hospitalization numbers that do not reflect the finalized values.

We queried these confirmed hospitalizations from the *covidData* package through the *covidHubUtils* package, both developed by Reich Lab at the University of Massachusetts Amherst to interface with COVID-19 surveillance data. Versions of data are stored such that users may access it as of a specified date [4]. For example, one may retrieve a version of the confirmed hospitalization dataset as of July 27, 2020. To ensure fair comparisons between the models developed for this analysis and some real-time forecasting models, our models trained on the confirmed hospitalizations that would have been available as of the “forecast date” of the retrospective forecast.

### 2.2 Forecasts

We define a forecast to be a quantitative prediction about the future based on already observed data, usually pertaining to a specific event, outcome, or trend related to one or more infectious diseases [13]. COVID-19 forecasting focuses on probabilistic forecasts because they quantify the likelihood of their occurrence, unlike simple point forecasts [8, 7, 13].

In this work, a forecast for a date-target-location combination (e.g. “1-day ahead COVID-19 incident hospitalizations in Massachusetts relative to 2020-07-04”) was described by one or both of a point forecast and a probabilistic forecast, represented by a set of 23 quantiles [8]. More information is given about forecast targets, horizons, locations, and quantiles later in this subsection.) According to conventions set by the Forecast Hub, all forecast values were truncated to be non-negative, as there is no real-world meaning for negative incident hospitalizations [5].

Further, while this was a retrospective analysis, forecasts were made with weekly Sunday forecast dates (to mirror how they would have been, if made in real-time) using the data available as of that day.

#### 2.2.1 Forecast horizons

Short-term forecasts for COVID-19 were defined to encapsulate horizons of 1-to 28-days ahead [8]. Forecasts for incident hospitalizations were made on a daily time scale, so we chose to restrict the forecast horizons we evaluated to 1-to 28-days ahead^1^. Horizons are related to target end dates in that a target end date of a particular forecast may be calculated by adding its horizon to the forecast date.

### 2.2.2 Forecast locations

We made forecasts for 53 of 55 locations from the HHS Protect confirmed hospitalizations dataset during the period of analysis. These 33 locations included the 30 states, two jurisdictions/territories Washington D.C. and Puerto Rico), and a U.S. national location. American Samoa and Guam were excluded due to their very low or zero COVID-19 incident hospitalizations over the entire period of analysis. Forecasts for low-count locations that predicted single-digit or zero hospitalizations were often very accurate; hence, these locations would not offer meaningful contributions to our understanding of differences in forecast skill between modeling approaches [8]. Further, we chose to evaluate the U.S. national location separately from the remaining 52 locations. This decision was motivated by the large difference in magnitude of incident hospitalizations between the two geographic scales, which would lead any unweighted averages of forecast accuracy metrics to favor models with high accuracy at the national level.

#### 2.2.3 Forecast quantiles

The Forecast Hub specified the usage of 23 quantiles

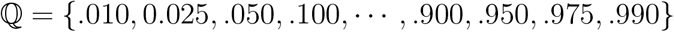

to represent a full probabilistic distribution, with the median 0.500 quantile usually taken as the model s point forecast. We followed this convention, though some additional work was needed to obtain probabilistic forecasts from the THieF methodology see subsection 2.3.4).

### 2.3 Date range of analysis

Ve focused on the period between Monday, July 27, 2020 and Sunday, October 2, 2022. This date range spans 798 days, or 114 weeks, which is just over 26 months. The entire period of analysis was split into two main phases: a validation phase of 462 days (66 weeks) and a testing phase of 236 days (48 weeks). The split between these two phases had the validation phase end on Sunday, October 31, 2021 and the testing phase begin on Monday, November 1, 2021.

The first day of the period of analysis (July 27, 2020) was selected as the first day of reliable truth data for all locations of interest. Forecasting for the validation phase began on Monday, December 7, 2020, as this was the first Monday after the HHS Protect confirmed hospitalizations dataset was declared the official ground truth data. The division between the validation and testing phases was chosen to encapsulate three COVID-19 variant-based waves in the validation phase (Winter 2020-21, Alpha, and Delta) and two waves in the testing phase (Omicron and BA.4/BA.5). The separation between pandemic waves was made based on U.S. national level incident hospitalizations, pictured below in Figure 1, and is generally consistent with splits made based on U.S. national incident cases or incident deaths.

**Figure 1:**
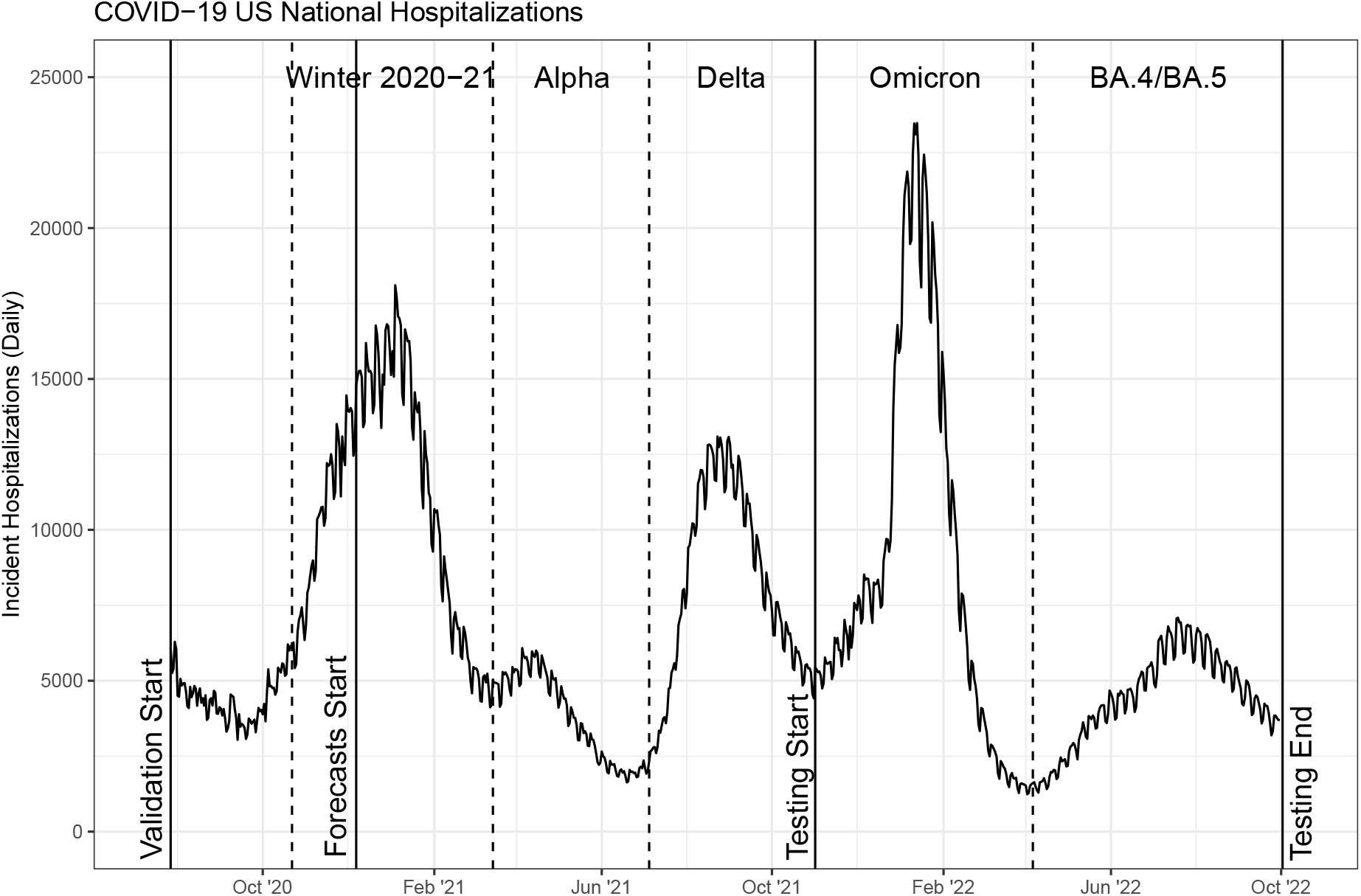
A plot of the period of analysis, superimposed over COVID-19 incident hospitalizations for the entire United States (U.S. national geographic scale). Vertical solid lines indicate divisions between phases while dashed lines indicate divisions between variant waves.

### 2.4 Benchmark comparison models

Six models from the Forecast Hub to serve as benchmark comparisons for both the validation and testing phases: four operational models, a naive baseline, and an equally-weighted median ensemble of all submitting incident hospitalization models. Forecasts from these models were queried from the Zoltar forecast archive, a research data repository developed by Reich Lab, through the *covidHub Utils* package [4, 23].

The four operational models consist of COVID-19 incident hospitalization models that submitted forecasts for all required locations, quantiles, and horizons as well as at least 90% of the forecast dates within the period of interest. Any models with systemic missingness that may prevent them from serving as useful benchmarks were also removed. (For example, Karlen-pypm did not submit predictions for most of the Omicron wave, a time during which making accurate forecasts was particularly difficult.) We also restricted the selection criteria to allow for a single model per submitting team. These models-JHUAPL-Bucky, USC-SI_kJalpha, CU-select, and GT-DeepCOVID-use mechanistic or machine learning methods when generating their predictions [7].

The COVIDhub-baseline is a naive baseline model whose forecasts are made using the previous day s incident hospitalization value as the point forecast (and 0.500 quantile for the probabilistic forecast) and past changes in daily incidence to calibrate uncertainty in its probabilistic forecasts [8]. Beating the baseline is typically seen as one marker of a useful model. The COVIDhub-4_week_ensemble is an equally-weighted median ensemble of eligible submitting models for incident hospitalizations at the COVID-19 Forecast Hub that makes short-term forecasts for 1-to 28 days ahead [5].

### 2.5 Temporal hierarchical forecasting (THieF)

Historically, hierarchical forecasting has been utilized in business, economics, operations management, and similar fields. This type of forecasting brings advantages like producing coherent forecasts^2^ and increasing prediction accuracy compared to other methods [1, 14]. Vhile most hierarchical forecasting considers cross-sectional hierarchies, not temporal ones, many of the same concepts may be applied to both types of hierarchies.

Forecast combination that integrates forecasts from different, independent modeling frameworks to create ensembles often results in better infectious disease forecasts compared to those from individual models [19, 18, 23, 3]. Likewise, classical statistical models like ARIMA models have demonstrated good performance for short-term forecasts in outbreak settings [15].

THieF offers a unique merging of hierarchical forecasting and forecasts combination of ARIMA models. Given the utility of its individual components, we believe that this method s success over more traditional hierarchical forecasting methods demonstrated in operations management [1] may carry over to predicting COVID-19 hospitalizations.

Surveillance data tends to exhibit noise due to reporting artifacts, especially when considering small time scales. The COVID-19 incident hospitalizations signal suffers a day-of-the-week effect as a result of hospital and clinic operating hours. Fewer hospitalizations are reported on Saturday and Sunday, instead often recorded as part of Monday hospitalizations, since not all facilities admit patients on weekends and those that do may not report those hospitalizations until the following week. We might better capture true short-term trends by aggregating daily hospitalizations to be weekly or every two weeks, although such smoothing carries some risk of excluding useful information at the daily level. Thus, we were careful to include both daily and weekly aggregation levels in the THieF temporal hierarchy construction for every model. Other higher aggregation levels were included in model schemes to investigate the existence of patterns at higher time scales and if such a pattern could improve model accuracy [12].

The THieF methodology consists of three steps to produce forecasts from timeseries data using a time-based hierarchical structure. These steps—data aggregation and hierarchy construction, base forecast creation, and reconciliation of base forecasts—are outlined in further detail below and in Figure 2 [1].

**Figure 2:**
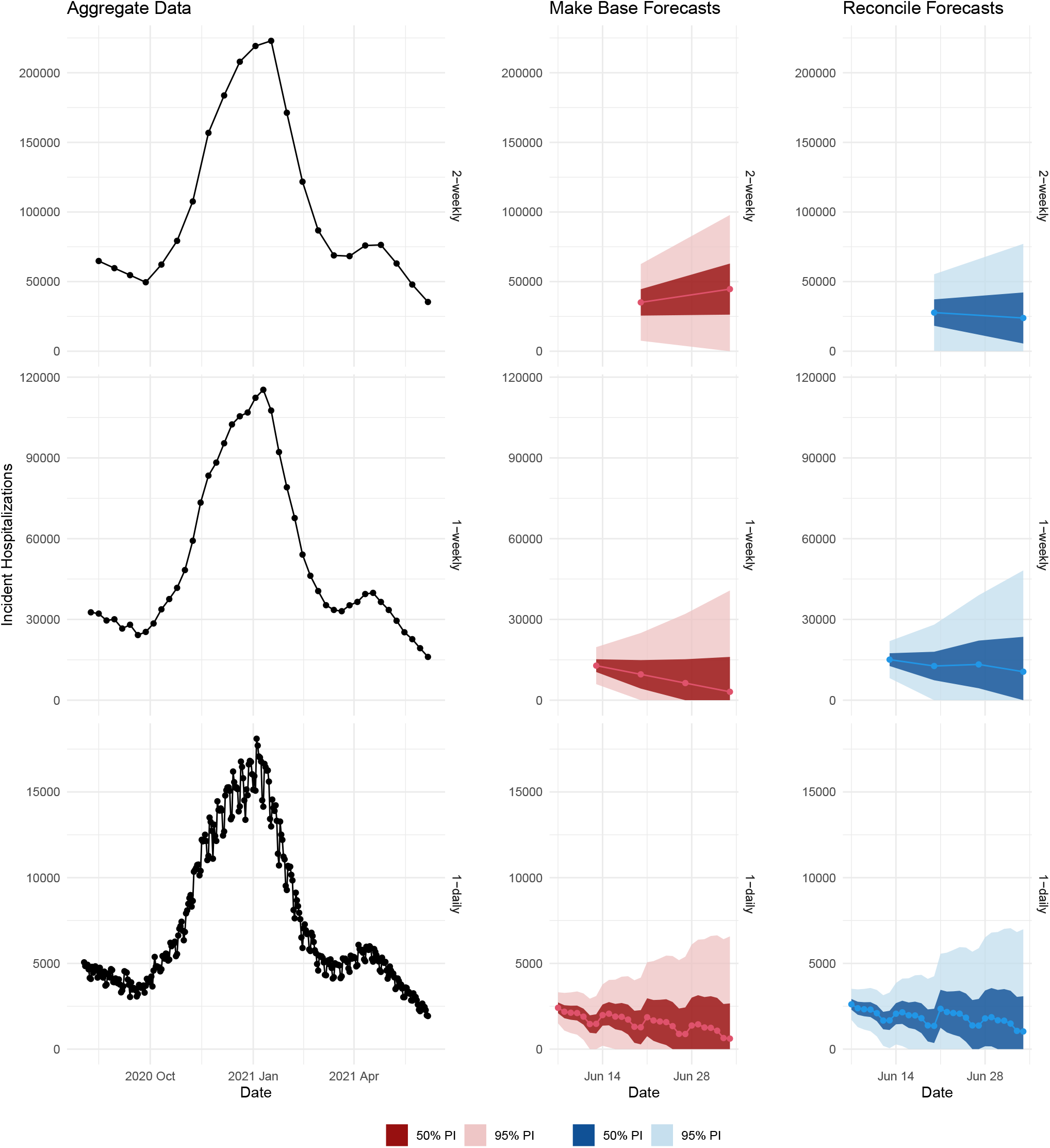
A plot showing the three steps of the THieF methodology for forecasting U.S. national incident hospitalizations for 1 to 28 days ahead on June 7, 2021

#### 2.5.1 Data Aggregation and Hierarchy

Suppose we wish to construct a THieF model that combines forecasts from 7- and 14-day aggregate time scales. We create calibration data for the hierarchy by summing the observed daily values over a particular timescale *k* [1]. By convention, we define all of our models to have a lowest and least-aggregate level to be that of the original time scale. Thus, our example model has three total levels: 1-day, 7-day, and 14-day see Figure 2). If desired, additional *k*-day levels can be added to this THieF model s temporal hierarchy as long as they are factors of the top-most aggregation level (14-days). Non-factor aggregation values will result in non-integer seasonal periods, and thus cannot be used [1]. A visual depiction of three possible THieF hierarchies is provided in Figure 9 in the Appendix.)

The incident hospitalizations for a particular location may be expressed generally as a time series {*y*_*t*_; *t* = 1, · · ·, *T*} when observed at the daily level. Let *m* be the number of observations of the smallest timescale (and the highest frequency) that aggregate to create a single data point for the most-aggregate level.(In our example *m* = 14.) These data are then aggregated to different *k*-day time scales based on the aggregation levels of interest for a particular model [1]. The variable *k* ∈ {*k*_*p*_, ·, *k*_2_, *k*_1_} can take on multiple values and indicates the particular aggregation level, where *k*_*p*_ = *m, k*_1_ = 1, and *p* is the total number of levels in the hierarchy [1]. In our example from earlier, *k* ∈ {14, 7, 1}, where *p* = 3.

Athanasopoulos, et al. propose an index *i* for each data point in the temporal hierarchy, defined based on the top, most-aggregate series 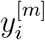 for *i* = 1, · · ·, ⌊*T/m*⌋.

A more general form for the time series for any level *k* is 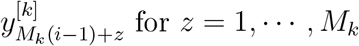 where *M*_*k*_ = *m/k* is the seasonal period for a particular level of *k*. An increase in the index *i* by 1 causes an increase of *M*_*k*_ periods for the time series at all aggregation levels, while an increase in *z* denotes a single period increase for the *k*-unit aggregation level [1].

For every index *i*, we construct *p* − 1 column vectors of dimension (*M*_*k*_ × 1)

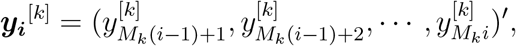

for all but the topmost level which only contains a single observation for each *i*.

These column vectors are then stacked into a larger (Σ_*k*_*M*_*k*_ × 1) column vector 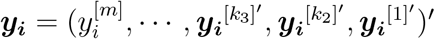 such that

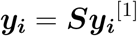

where ***S*** is the (Σ_*k*_*M*_*k*_ × *m*) summing matrix from [13] and ***y***_***i***_^[1]^ is the (*m* × 1) vector of daily observations with index *i*. For a hierarchy with levels *k* ∈ 14, 7, 1, this equation expands out to

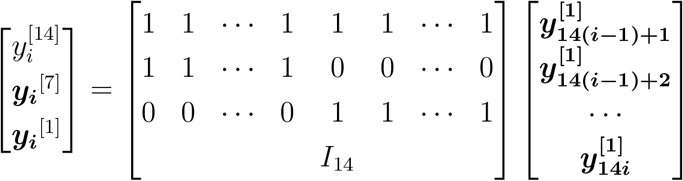

in which the dimensions of the matrices are (17×1), (17×14), and (14×1), respectively.

#### 2.5.2 Base Forecasts

Next, base forecasts are made at every level of the hierarchy, treating each aggregation level as independent univariate time series (see Figure 2). We fit an ARIMA model to each level of the series—which was found to produce the most accurate reconciled THieF forecasts—[1] and allow the parameters to vary for each level. At this stage, the base forecasts are usually incoherent, which may indicate that important patterns have been detected at different time scales using the THieF method.

The base forecasts have the form *ŷM*_*k*_ (*g*−1)+*z*, where *g* represents the forecast horizon and is based on the index for the top-level series. Like with the observed data, we then construct (Σ_*k*_*M*_*k*_ × 1) column vectors for each forecast horizon *g*:

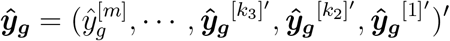

where each element 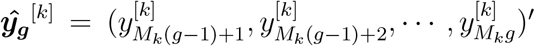 is a (*M*_*k*_ × 1) column vector of the base forecasts from that level [1].

The base forecasts may also be expressed as

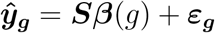

where ***S*** is the summing matrix defined in the previous subsection, 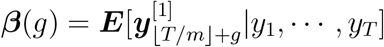 is the unknown mean of future daily observed values conditional on the currently observed data points; and ***ε***_***g***_ is the difference between the base forecasts ***ŷ***_*g*_ and the expected value of their corresponding reconciled forecasts, termed the reconciliation error, assuming that *ε*_*g*_ has zero mean and covariance matrix **Σ** [1].

#### 2.5.3 Reconciled Forecasts

Through the step of reconciliation, we integrate the information from all the aggregation levels into the output forecasts. Ideally, we want to combine the base forecasts ***ŷ***_***g***_ in a way that minimizes the difference between them and the reconciled forecasts 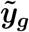. As the covariance matrix of **Σ** is non-identifiable 1, we must estimate **Σ** using a weighted least squares WLS) estimator **Λ**_***SV***_ called the series variance estimator to obtain the reconciled forecasts

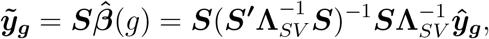

where ***S*** is the previously defined summing matrix and 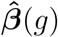 is an estimator of the unknown conditional mean ***β***(*g*) [1].

The series variance estimator is a diagonal (Σ_*k*_*M*_*k*_ × Σ_*k*_*M*_*k*_) matrix of weights proportional to the error variance of each aggregated series that make up the levels of the hierarchy. In other words, the series variance estimator assumes that variance is constant across the forecasts for each time-series, a common assumption for time-series data [1]. For the THieF model with the top-most 14-day aggregation level, this matrix is given by

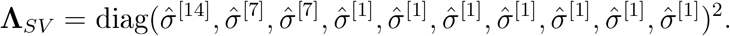

While Athanasopoulos, et al. proposed two other alternative WLS estimators, they later showed that **Λ**_*SV*_ yields the most accurate reconciled forecasts. All of these VLS estimators are estimators for the sample covariance estimator **Λ** for ***W***, the covariance matrix of the base forecast error. In turn, ***W*** is an estimator for the unknowable **Σ** [1].

The reason we prefer the series variants estimate rover the sample covariance estimator **Λ** is that calculation of the latter is difficult to implement in practice ^3^. This estimator has (Σ_*k*_*M*_*k*_)^2^ elements that must be estimated, and the sample size that can be used for such estimation is limited by the number of observations at the topmost aggregation level ⌊*T/m*⌋. Since ⌊*T/m*⌋ ≪ *T*, the accuracy of the estimation is degraded. Hence, we instead choose to rely on **Λ**_*SV*_ [1].

Following this final step of reconciliation, the forecasts from every time scale will agree about the number of daily incident hospitalizations for all horizons. The reconciled forecasts are shown in Figure 2.

#### 2.5.4 Making THieF Probabilistic

Recall that COVID-19 forecasts are probabilistic. However, the THieF methodology defined above and in [1] is used for point forecasting. To address this challenge, we calculated prediction intervals for the base forecasts, reconciled the endpoints as if they were point forecasts using the same weighted least squares procedure described in the previous subsection, then translated the prediction interval bounds into quantiles from a full probabilistic distribution. The translation is summarized in Table 1. For example, the values for a 0.500 and a 0.950 quantile (see the Quantile row) may be obtained by taking the lower and upper bounds of the 90% prediction interval (see the Interval row, where “L” and “U” represent the lower and upper bounds, respectively).

**Table 1:** Table of the 23 quantiles required by the Forecast Hub and their associated prediction interval bound.

|  |  |  |  |  |  |  |  |  |  |  |  |
| --- | --- | --- | --- | --- | --- | --- | --- | --- | --- | --- | --- |
| Quantile | .010 | .025 | .050 | 0.100 | ... | 0.500 | ... | 0.900 | 0.950 | .975 | .990 |
| Interval | 98L | 95L | 90L | 80L | ... | Median | ... | 80U | 90U | 95U | 98U |

Although this means that the interval forecasts are not coherent (only the point forecasts are) and it would be preferable to obtain quantiles from the reconciled probabilistic forecast distribution itself, we generally did not find a concerning drop in prediction interval coverage rates for our evaluations conducted during either the validation phase or the testing phase see subsection 3).

### 2.6 Model specifications

We created four distinct classes of models for comparison during the validation phase: the original THieF models, the THieF ensembles, the ARIMA and SARIMA models, and an untrained mean ensemble of the ARIMA and SARIMA models. In total, these four groups consisted of twenty-seven (27) candidate models, all of which were implemented using the forecast package in R.

#### 2.6.1 Original THieF models

The original THieF models were constructed using the THieF methodology outlined in subsection 2.3 with different combinations of aggregation levels and data transformations. Seven hierarchical structures and two data transformations resulted in fourteen (14) original THieF models. Each model was defined (and named) based on the highest level of its temporal hierarchy and the data transformation that is used before generating the forecasts. For example, the simplest of the THieF models is THieF_1wk-noTransform while the most complex is THieF_12wk-4root.

The aggregation levels of any model s temporal hierarchy were daily, plus *f*_*m*_-week periods where *f*_*m*_ is the set of all factors for the highest time scale *m*. Hence, the THieF_12wk-4root model (as well as the THieF_12wk-noTransform model) had a temporal hierarchy made up of daily, weekly, 2-weekly, 3-weekly, 4-weekly, 6-weekly, and 12-weekly aggregation levels since *f*_12_ = {1, 2, 3, 4, 6, 12}. Meanwhile, the THieF_1wk-noTransform model s temporal hierarchy was only made up of daily and weekly aggregation levels (*f*_1_ = {1}). We chose 1-week, 2-week, 3-week, 4-week, 6-week, 8-week, and 12-week top-level aggregations so as to have a range of time scales to inform the reconciled forecasts. Supplemental Figure 9 illustrates the construction of several models temporal hierarchies.

We explored these seven hierarchy schemes with both no data transformation and a fourth-root data transformation. The latter transform was implemented by first taking the fourth root of the confirmed hospitalizations time series, using the truth data to make forecasts with the THieF methodology, then performing an inverse fourth-power transformation to the resulting forecasts. We selected the fourth-root transformation over more common log or square root transforms based on prior research showing accuracy improvements for COVID-19 forecasting due to its variance-stabilizing effect [21].

#### 2.6.2 THieF ensemble models

The THieF ensemble models, defined using equations adapted from Ray, et al. [18], consisted of eight mean ensembles of the fourteen total original THieF models. Vhile not all fourteen original THieF models were passed onto the testing phase, the THieF ensembles that were selected remain ensembles of these fourteen original THieF models (see subsection 2.7.2).

Of the eight THieF ensembles, we formulated seven as trained ensembles, in which past performance of the component THieF models was used to assign the components weights, and one as an untrained ensemble, in which all component forecasters were assigned equal weight [18]. Both types of ensembles made quantile forecasts^4^ at level *u* by combining those of the component forecasts for a particular forecast date, location, horizon, and quantile combination. An ensemble quantile forecast can be expressed mathematically as

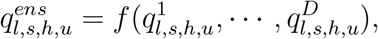

where *l* is the location, *s* is the forecast date, *h* is the horizon, and the superscript *d* = 1, · · ·, *D* denotes the model [18].

A trained, weighted mean ensemble calculated its forecast quantiles as

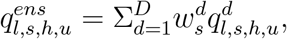

in which the weights were calculated using a sigmoidal transformation of the component models relative WIS (rWIS) over a rolling window of twelve (12) weeks leading up to the ensemble s forecast date *s*:

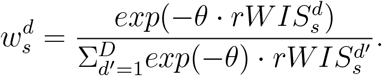

When the non-negative parameter *θ* = 0, this became an equally weighted ensemble. However, as *θ* increases, the weights became more unequal, with the contributions of better-performing component models eclipsing that of worse-performing models. Note that the weights, however, were always non-negative and summed to one. A group of weights was calculated for a particular forecast date, location, horizon, and quantile combination [18].

The THieF ensembles were named with the *θ* value used in their construction, except for the untrained ensemble, which was called THieF_ensemble-mean. We selected *θ* values {0, 1, 3, 6.3, 10, 13, 20, 23} to encompass the range of values taken on by the COVIDhub-trained_ensemble^5^ when predicting incident hospitalization [18]. The one non-integer value 6.3) was selected as the median (and approximate mean) *θ* value over the period of analysis for all locations of interest except for the U.S. national level. The corresponding ensemble was named THieF_ensemble-train6.5.

While the value of *θ* generally remained static for each ensemble, all trained ensembles were defined to start with *θ* = 0 until 12 weeks of prior forecasts were achieved on March 1, 2021) during the validation phase that could be used to calculate weights based on prior performance of forecasts. During the testing phase, the value of the weights was dependent on at least some validation phase performance until the thirteenth week beginning on January 24, 2022).

#### 2.6.3 ARIMA and SARIMA models

The four ARIMA and SARIMA (seasonal ARIMA) models were classic statistical models created using the *auto arma ()* function from the forecast package in R. This function makes forecasts based on input time series data, and the user may specify seasonality when creating a SARIMA model. Using the original HHS Protect confirmed hospitalizations dataset, we created two ARIMA models and two SARIMA models, with either no data transformation or a variance-stabilizing fourth root transformation, as described in subsection 2.6.1. The SARIMA models both had a seasonality of 7 days 1 week), named sarima_s7-4root and sarima_s7-noTransform, while the ARIMA models had no seasonality, named arima_s1-4root and arima_s1-noTransform.

The construction of the ARIMA and SARIMA models intentionally matched that of the daily and weekly aggregation levels of the original THieF models, though in practice the resulting base forecasts from the THieF models may have differed slightly due to the aggregation process to create the temporal hierarchy. We specifically wanted to use the ARIMA and SARIMA models to help investigate whether the THieF methodology s temporal hierarchy and forecast reconciliation impacted its performance rather than simply its underlying ARIMA base forecaster. (Recall that research has shown that classic statistical models have shown good performance for outbreak forecasting, thus they were chosen as reasonable comparators [15].)

#### 2.6.4 ARIMA and SARIMA ensemble

The ARIMA and SARIMA ensemble, named sarima-untrained_ensemble, is an untrained quantile mean of all the models described in the previous subsection. We formulated this ensemble to investigate whether the source of any potential performance improvements with THieF are a result of just combining multiple ARIMA forecasts, or the result of weighted, temporal reconciliation of ARIMA forecasts at different time scales.

### 2.7 Metrics and Evaluation

Evaluation of our models was conducted by scoring the forecasts for every unique combination of model, location, horizon, and forecast date using several metrics of interests, then grouping together the scores and taking averages of these metrics over various aggregations (described in more detail later in this subsection). We used the same metrics as the Forecast Hub to for both the validation and testing phases: average mean absolute error (average MAE) for point forecasts, and average weighted interval score (average WIS, taken from 2), average 50% prediction interval PI) coverage, and average 95% PI coverage for probabilistic forecasts. (We refer to these metrics simply as MAE, WIS, and 50% and 95% PI coverage going forward for simplicity.) For WIS and MAE, lower values are desirable while coverage levels approximating 1 − *α* indicate a well-calibrated forecast. This means that the intervals were narrow enough to provide useful information but not so narrow as to exclude the truth.

Averages for the four metrics were first calculated for probabilistic forecasts (and point forecasts) for a single forecast date, location, horizon, and quantile combination, then the mean was taken for each model from every forecast aggregation. These calculations were performed using the following equations, which are adapted from [8] :

1. 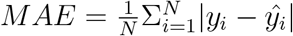
2. 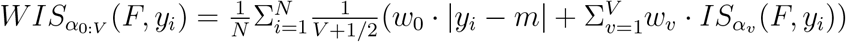
3. (1 − *α*) * 100%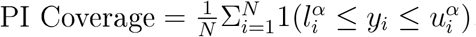,

where *N* is the number of forecasts being averaged, *y*_*i*_ is the true observed value at a particular time point (date) and location, and *ŷ*_*i*_ is the point forecast value for a particular forecast date, horizon, and location combination. The PI uncertainty level is given by *α, v* is the particular prediction interval (*v* = 1, · · ·, *V*), and *F* is the forecast distribution. The weights *w*_0_ and *w*_*v*_ were set as *w*_0_ = 1*/*2 and *w*_*v*_ = *α*_*v*_*/*2 respectively while 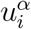 and 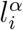 are the upper and lower bounds of a (1 − *α*) * 100%-level prediction interval for a particular forecast date and location. Lastly, the interval score of a particular (1 − *α*) * 100%-level prediction interval is 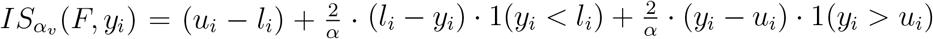 where 1() is the indicator function [8].

Weighted interval score is a proper scoring rule for a set of interval forecasts comprised of three penalty components—spread, overprediction, and underprediction—that measures how closely a set of prediction intervals is consistent with the true observed value. WIS is an alternative to more common proper scoring rules like the logarithmic score (logS) and continuous ranked probability score (CRPS) which can not be evaluated directly for interval forecasts. However, with the weights *w*_0_, · · ·, *w*_*V*_ set as described above, large *V*, and equally spaced values of *α*_1_, · · ·, *α*_*V*_, it can be shown that 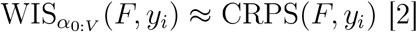

Relative versions of MAE and WIS were also used to adjust for truth data magnitude to allow for comparison between forecast values of different scales. These relative metrics were calculated as *MAE*_*model d*_ */MAE*_*baseline*_ and *WIS*_*model d*_ */WIS*_*baseline*_, respectively [8].

The scores were then aggregated in one of seven ways: overall validation phase, last 17 weeks of the validation phase, overall testing phase, by pandemic wave for the testing phase, by forecast date for the testing phase, and by location for the testing phase. We separate the evaluations of the U.S. national location from the other 52 locations to avoid inflated scores that disproportionately represent a single location, as scores are generally higher when counts are higher and counts are higher for larger population locations. Additionally, the by forecast date aggregation of scores was performed weekly instead of daily, following conventions set by the Forecast Hub for evaluation reports to condense the amount of scores. These weekly scores were obtained by averaging score values across a single week (Tuesday through Monday) [6] ^6^.

We performed no statistical tests to assess the significance of performance differences between models, as certain issues arise when making time series forecasts that prevent usage of such tests. Namely, scores tend to be highly correlated across multiple dimensions, like horizons, forecast dates, locations, etc. Assumptions of independent, identically distributed data found in standard inference are invalid in time series forecasting. In practice, numerical comparisons of forecast error between prospective models without excluding any models that were compared) is sufficient for choosing the best model to inform decision-making. Additionally, foregoing formal statistical tests is common practice in infectious disease forecasting. Still, it should be noted that we cannot necessarily make a definitive statement about the predominance of one model over another without such tests [15].

#### 2.7.1 Validation phase

The validation phase is used to train the candidate models, calibrate the forecasts, and to select the best models to pass on to the testing phase based on their performance. All validation phase evaluations are restricted to forecasts with target end dates (date for which the prediction of incident hospitalizations is being made) within the validation phase (July 27, 2020 to October 31, 2021), with forecasts beginning on Monday, December 7, 2020.

We prespecified a set of criteria to investigate several objectives of interest to help us determine which models to pass along as part of the best subset:

- **What is the best model created using THieF over the entire validation phase?** Best THieF model (not including ensembles) over the entire validation phase
- **Does ensembling THieF models with accuracy-based weights improve performance?** Best trained ensemble of THieF models over the entire validation phase
- **Does THieF improve upon performance compared to its base forecaster?** Best ARIMA/SARIMA model over the entire validation phase
- **Does THieF improve upon performance compared to an ensemble of its base forecaster?** ARIMA and SARIMA model ensemble
- **How does a simple equally-weighted ensemble compare against trained ensembles?** Untrained ensemble of THieF models
- **Do some THieF models improve with more input data?** Best model created using THieF during the last 17 weeks of the validation phase

where “best” is defined by lowest WIS. These criteria involve passing on a representative from each class of models—original THieF, THieF (trained) ensemble, ARIMA/SARIMA-plus the equally weighted THieF ensemble and an additional original THieF model based on performance during the last 17 weeks of the validation phase. We limit the last category to original THieF models because we suspected those with large aggregation levels might fit this description. Additionally, we allow each geographic scale to put forth a model for each category when applicable. Thus, a maximum of ten models could make up the best subset to move onto the testing phase evaluations, if unique candidates occupied each category for each geographic scale. However, if the same model occupied multiple categories for both states and U.S. national, we might pass on only five models.

#### 2.7.2 Testing phase

Ve use the best subset to make forecasts during the testing phase and evaluated model accuracy among this subset and against the six comparison models from the Forecast Hub: COVIDhub-baseline and COVIDhub-4_week_ensemble and the four operational models. Like with the validation phase, all testing phase evaluations are restricted to forecasts with target end dates within the testing phase (November 1, 2021 to October 2, 2022).

The testing phase analysis is broken down into five smaller evaluations, based on aggregating the testing phase forecasts for the overall period, by horizon week, by pandemic wave, by location, and by forecast date. The smaller evaluations are intended to help pinpoint any potential accuracy gains (or losses) the original THieF or THieF ensembles had over the COVIDhub-baseline (or COVIDhub-4_week_ensemble) and allow us to hypothesize the origin of these accuracy differences.

The overall period evaluation provides a generalized view of the testing phase analysis and is meant to identify a single best model that could be used to inform decision-making. The by-horizon evaluation helps identify models that perform well at shorter horizons typically (1 to 2 week ahead), as the larger WIS and MAE values for longer horizons 3 to 4 week ahead) likely dominated these metrics in the overall period evaluation [20].

The by–pandemic wave and by forecast date evaluations investigate time periods with different magnitudes or fluctuating rates of incident hospitalizations. Times of greater values or most rapid changes have larger WIS and MAE and are generally harder to predict [10, 20], and it might be useful to identify which models performed best under different scenarios. These two evaluations are also stratified by horizon week. However, the by-location evaluation, which provided insight into differences in model performance between higher and lower count locations, is not stratified by horizon week.

### 2.8 Software and Reproducibility

Code for the manuscript and supplement can be found at https://github.com/lshandross/covidTHieF. Functions for making the forecasts are based on the TH eF package. The manuscript is generated with a reproducible workflow using the arge s package. All analysis was performed using R version 4.4.1 [17].

## 3 Results

### 3.1 Validation phase

During the validation phase, we found that the THieF ensembles generally made the most accurate predictions of incident COVID-19 hospitalizations of all our constructed models, followed by the ARIMA/SARIMA ensemble, then the original THieF models, and finally the ARIMA/SARIMA models. These results are summarized in Tables 2 and 3, which shows the overall model ranking and evaluation metrics ordered by ascending WIS. A complete list of the best subset models passed onto the testing phase is provided in the Appendix in Table 6.

**Table 2:** Summary of overall model performance during the validation phase for averaged states, ordered by ascending WIS. Models part of the best subset are denoted with an asterisk *). The COVIDhub-baseline is included as a point of reference.

|  | Model | WIS | MAE | Cov50 | Cov95 | rWIS | rMAE |  |
| --- | --- | --- | --- | --- | --- | --- | --- | --- |
| 1 | THieF_ensemble-train10 | 30.73 | 47.22 | 0.53 | 0.93 | 0.86 | 0.94 |  |
| 2 | THieF_ensemble-train15 | 30.74 | 47.28 | 0.52 | 0.93 | 0.86 | 0.95 |  |
| 3 | THieF_ensemble-train6.5 | 30.77 | 47.20 | 0.53 | 0.94 | 0.86 | 0.94 |  |
| 4 | THieF_ensemble-train20 | 30.77 | 47.33 | 0.52 | 0.93 | 0.86 | 0.95 |  |
| 5 | THieF_ensemble-train25 | 30.81 | 47.37 | 0.51 | 0.93 | 0.86 | 0.95 |  |
| 6 | THieF_ensemble-train3 | 30.90 | 47.25 | 0.54 | 0.94 | 0.86 | 0.95 | * |
| 7 | THieF_ensemble-train1 | 31.33 | 47.44 | 0.53 | 0.94 | 0.87 | 0.95 |  |
| 8 | THieF_6wk-4root | 31.65 | 48.11 | 0.46 | 0.91 | 0.88 | 0.96 | * |
| 9 | THieF_6wk-noTransform | 32.26 | 48.67 | 0.54 | 0.92 | 0.90 | 0.97 |  |
| 10 | sarima-untrained_ensemble | 32.46 | 49.06 | 0.54 | 0.91 | 0.90 | 0.98 | * |
| 11 | THieF_1wk-4root | 32.68 | 50.06 | 0.48 | 0.91 | 0.91 | 1.00 |  |
| 12 | THieF_2wk-4root | 32.94 | 50.26 | 0.47 | 0.91 | 0.92 | 1.01 |  |
| 13 | THieF_2wk-noTransform | 33.26 | 48.03 | 0.57 | 0.90 | 0.93 | 0.96 |  |
| 14 | THieF_1wk-noTransform | 33.74 | 49.21 | 0.58 | 0.90 | 0.94 | 0.98 |  |
| 15 | THieF_8wk-4root | 33.84 | 51.14 | 0.43 | 0.89 | 0.94 | 1.02 |  |
| 16 | THieF_4wk-4root | 34.10 | 51.50 | 0.46 | 0.90 | 0.95 | 1.03 |  |
| 17 | THieF_ensemble-mean | 34.29 | 47.65 | 0.53 | 0.94 | 0.96 | 0.95 | * |
| 18 | THieF_3wk-4root | 34.62 | 52.71 | 0.46 | 0.90 | 0.96 | 1.05 |  |
| 19 | THieF_8wk-noTransform | 34.76 | 52.06 | 0.51 | 0.90 | 0.97 | 1.04 |  |
| 20 | THieF_3wk-noTransform | 34.96 | 52.08 | 0.54 | 0.90 | 0.97 | 1.04 |  |
| 21 | THieF_4wk-noTransform | 35.63 | 52.82 | 0.53 | 0.89 | 0.99 | 1.06 |  |
| 22 | COVIDhub-baseline | 35.88 | 49.98 | 0.83 | 0.98 | 1.00 | 1.00 |  |
| 23 | sarima_s7-noTransform | 36.29 | 51.91 | 0.57 | 0.89 | 1.01 | 1.04 | * |
| 24 | sarima_s7-4root | 36.35 | 54.55 | 0.46 | 0.90 | 1.01 | 1.09 |  |
| 25 | arima_s1-4root | 36.91 | 55.43 | 0.46 | 0.89 | 1.03 | 1.11 |  |
| 26 | THieF_12wk-noTransform | 39.14 | 59.06 | 0.42 | 0.89 | 1.09 | 1.18 |  |
| 27 | arima_s1-noTransform | 39.41 | 56.38 | 0.54 | 0.88 | 1.10 | 1.13 |  |
| 28 | THieF_12wk-4root | 84.32 | 54.49 | 0.41 | 0.86 | 2.35 | 1.09 | * |

**Table 3:** Summary of overall model performance during the validation phase for the entire U.S., ordered by ascending WIS. Models part of the best subset are denoted with an asterisk (*). The COVIDhub-baseline is included as a point of reference.

|  | Model | WIS | MAE | Cov50 | Cov95 | rWIS | rMAE |  |
| --- | --- | --- | --- | --- | --- | --- | --- | --- |
| 1 | sarima-untrained_ensemble | 1104.9 | 1778.1 | 0.30 | 0.86 | 0.87 | 0.89 | * |
| 2 | THieF_6wk-noTransform | 1116.4 | 1756.0 | 0.36 | 0.91 | 0.88 | 0.88 | * |
| 3 | THieF_ensemble-train3 | 1139.8 | 1808.6 | 0.34 | 0.91 | 0.89 | 0.91 | * |
| 4 | THieF_ensemble-train1 | 1144.9 | 1814.9 | 0.35 | 0.90 | 0.90 | 0.91 |  |
| 5 | THieF_ensemble-train6.5 | 1146.6 | 1817.4 | 0.34 | 0.91 | 0.90 | 0.91 |  |
| 6 | THieF_6wk-4root | 1152.4 | 1765.8 | 0.33 | 0.76 | 0.90 | 0.88 |  |
| 7 | THieF_ensemble-train10 | 1159.2 | 1834.0 | 0.33 | 0.91 | 0.91 | 0.92 |  |
| 8 | THieF_ensemble-train15 | 1180.2 | 1866.3 | 0.33 | 0.90 | 0.92 | 0.94 |  |
| 9 | THieF_ensemble-train20 | 1199.4 | 1897.5 | 0.32 | 0.90 | 0.94 | 0.95 |  |
| 10 | THieF_ensemble-mean | 1212.7 | 1822.3 | 0.35 | 0.90 | 0.95 | 0.91 | * |
| 11 | THieF_ensemble-train25 | 1214.3 | 1919.3 | 0.32 | 0.89 | 0.95 | 0.96 |  |
| 12 | THieF_1wk-noTransform | 1218.5 | 1856.4 | 0.46 | 0.87 | 0.95 | 0.93 |  |
| 13 | THieF_2wk-4root | 1234.7 | 1941.8 | 0.25 | 0.85 | 0.97 | 0.97 |  |
| 14 | THieF_1wk-4root | 1252.2 | 1993.1 | 0.30 | 0.85 | 0.98 | 1.00 |  |
| 15 | COVIDhub-baseline | 1276.5 | 1995.9 | 0.64 | 0.99 | 1.00 | 1.00 |  |
| 16 | THieF_8wk-noTransform | 1277.0 | 1968.9 | 0.38 | 0.85 | 1.00 | 0.99 |  |
| 17 | THieF_8wk-4root | 1303.6 | 1963.8 | 0.26 | 0.70 | 1.02 | 0.98 |  |
| 18 | THieF_2wk-noTransform | 1343.1 | 1985.2 | 0.40 | 0.82 | 1.05 | 0.99 |  |
| 19 | THieF_4wk-4root | 1383.4 | 2058.0 | 0.28 | 0.70 | 1.08 | 1.03 |  |
| 20 | THieF_3wk-noTransform | 1392.9 | 2008.6 | 0.38 | 0.78 | 1.09 | 1.01 |  |
| 21 | THieF_4wk-noTransform | 1397.4 | 2038.4 | 0.35 | 0.77 | 1.09 | 1.02 |  |
| 22 | THieF_3wk-4root | 1401.7 | 2164.8 | 0.26 | 0.80 | 1.10 | 1.08 |  |
| 23 | arima_s1-noTransform | 1477.9 | 2162.0 | 0.37 | 0.79 | 1.16 | 1.08 | * |
| 24 | sarima_s7-noTransform | 1483.2 | 2200.6 | 0.39 | 0.81 | 1.16 | 1.10 |  |
| 25 | arima_s1-4root | 1530.2 | 2227.1 | 0.26 | 0.75 | 1.20 | 1.12 |  |
| 26 | THieF_12wk-noTransform | 1553.5 | 2357.8 | 0.32 | 0.76 | 1.22 | 1.18 |  |
| 27 | sarima_s7-4root | 1601.5 | 2467.2 | 0.22 | 0.76 | 1.25 | 1.24 |  |
| 28 | THieF_12wk-4root | 2270.6 | 2039.9 | 0.34 | 0.66 | 1.78 | 1.02 | * |

### 3.2 Testing phase

The eight models from the best subset were joined by the six Forecast Hub comparison models for the testing phase analysis. Generally, we found that the COVIDHub-4_week_ensemble, CU-select, and the original THieF models with a fourth root transformation showed the best performance out of the fourteen total models across all of the smaller evaluations. The THieF ensembles displayed more middling performance, though they almost always beat the COVIDhub-baseline.

Figure 3 shows 1-to 28-day ahead forecasts during the testing phase for a top performing model (THieF_6wk-4root), a poor performing model (sarima_s7-noTransform), the COVIDhub-baseline, and the COVIDhub-4_week_ensemble. Plots of additional forecasts for a subset of locations made by every model compared during the testing face can be found in the Supplemental Forecast Plots file. The COVIDhub-4_week_ensemble followed the confirmed hospitalizations better most other models, especially during the Omicron wave s period of most rapid increase; its coverage rates also generally achieved or even exceeded the nominal level. CU-select also showed great adherence to the observed hospitalization values but demonstrated very low coverage rates. Except for times a rapid change during the Omicron wave, the THieF models displayed a good balance of matching the observed data and have a well-calibrated forecast intervals. The COVIDhub-baseline generally had the widest 95% interval forecasts of the plotted models. Lastly, the other operational models struggled to follow the trajectory of COVID-19 hospitalizations and faced some calibration-related issues. These results are mirrored in the subsequent stratified evaluations.

**Figure 3:**
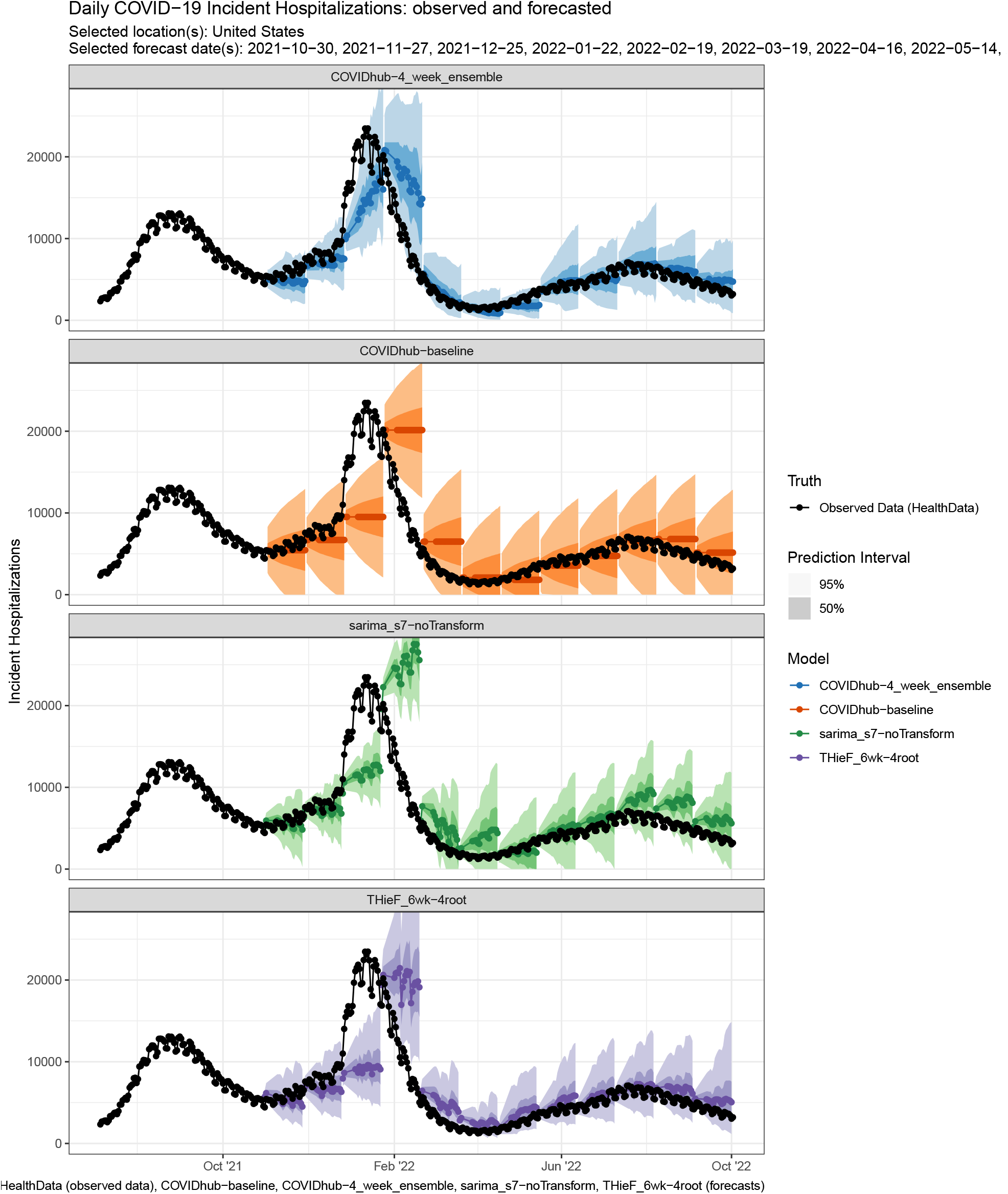
Quantile forecasts for daily incident COVID-19 hospitalizations in the entire U.S. during the testing phase from the COVIDhub-4_week_ensemble, the COVIDhub-baseline, a poor performing model (sarima_s7-noTransform), and a top performing model (THieF_6wk-4root). Forecasts are represented by a (median) point forecast, 30% and 90% prediction intervals.

#### 3.2.1 Overall model performance

Model rankings were generally consistent between the averaged states and U.S. national scales for the overall testing phase evaluation, with the top seven or eight models outperforming the COVIDhub-baseline (Tables 4 and 3). The COVIDhub-4_week_ensemble has the lowest WIS and MAE by a substantial margin, followed by THieF_6wk-4root or CU-select, THieF_12wk-4root, THieF_ensemble-train3,and THieF_ensemble-mean. The models that performed worse than the baseline tended to be consistent between between geographic scales but had more fluctuating rankings. However, the coverage rates differed for the two location-based groupings, with models tending to display higher coverage rates for the averaged states scale compared to the U.S. national one. In particular, CU-select had some of the lowest coverage rates for a top-performing model (Tables 4 and 3).

**Table 4:** Summary of overall model performance during the testing phase for averaged states, ordered by ascending WIS.

|  | Model | WIS | MAE | Cov50 | Cov95 | rWIS | rMAE |
| --- | --- | --- | --- | --- | --- | --- | --- |
| 1 | COVIDhub-4_week_ensemble | 30.18 | 44.87 | 0.64 | 0.94 | 0.72 | 0.79 |
| 2 | THieF_6wk-4root | 34.55 | 50.12 | 0.51 | 0.90 | 0.83 | 0.88 |
| 3 | CU-select | 34.89 | 49.98 | 0.32 | 0.75 | 0.84 | 0.88 |
| 4 | THieF_12wk-4root | 35.25 | 51.35 | 0.50 | 0.89 | 0.85 | 0.90 |
| 5 | THieF_ensemble-train3 | 35.62 | 51.57 | 0.56 | 0.91 | 0.86 | 0.91 |
| 6 | THieF_ensemble-mean | 35.91 | 51.80 | 0.57 | 0.91 | 0.86 | 0.91 |
| 7 | THieF_6wk-noTransform | 39.51 | 56.87 | 0.55 | 0.88 | 0.95 | 1.00 |
| 8 | COVIDhub-baseline | 41.63 | 56.77 | 0.79 | 0.96 | 1.00 | 1.00 |
| 9 | sarima-untrained_ensemble | 42.06 | 56.87 | 0.60 | 0.86 | 1.01 | 1.00 |
| 10 | arima_s1-noTransform | 42.92 | 58.04 | 0.60 | 0.86 | 1.03 | 1.02 |
| 11 | sarima_s7-noTransform | 43.54 | 58.94 | 0.60 | 0.87 | 1.05 | 1.04 |
| 12 | GT-DeepCOVID | 44.16 | 57.82 | 0.28 | 0.59 | 1.06 | 1.02 |
| 13 | JHUAPL-Bucky | 44.67 | 65.69 | 0.48 | 0.89 | 1.07 | 1.16 |
| 14 | USC-SI_kJalpha | 70.83 | 90.52 | 0.21 | 0.50 | 1.70 | 1.59 |

**Table 5:** Summary of overall model performance during the testing phase for the entire U.S., ordered by ascending WIS.

|  | Model | WIS | MAE | Cov50 | Cov95 | rWIS | rMAE |
| --- | --- | --- | --- | --- | --- | --- | --- |
| 1 | COVIDhub-4_week_ensemble | 1401.3 | 2085.1 | 0.63 | 0.91 | 0.75 | 0.83 |
| 2 | CU-select | 1479.2 | 2186.3 | 0.28 | 0.71 | 0.79 | 0.87 |
| 3 | THieF_6wk-4root | 1642.5 | 2358.8 | 0.40 | 0.82 | 0.88 | 0.94 |
| 4 | THieF_ensemble-train3 | 1687.4 | 2443.4 | 0.36 | 0.83 | 0.90 | 0.97 |
| 5 | THieF_ensemble-mean | 1700.5 | 2458.9 | 0.39 | 0.84 | 0.91 | 0.98 |
| 6 | THieF_12wk-4root | 1723.9 | 2481.7 | 0.31 | 0.81 | 0.92 | 0.99 |
| 7 | THieF_6wk-noTransform | 1727.7 | 2504.8 | 0.46 | 0.86 | 0.92 | 1.00 |
| 8 | GT-DeepCOVID | 1864.9 | 2455.9 | 0.26 | 0.55 | 1.00 | 0.98 |
| 9 | COVIDhub-baseline | 1867.5 | 2514.1 | 0.71 | 0.85 | 1.00 | 1.00 |
| 10 | JHUAPL-Bucky | 1888.7 | 2707.8 | 0.69 | 0.97 | 1.01 | 1.08 |
| 11 | arima_s1-noTransform | 1949.6 | 2487.0 | 0.53 | 0.82 | 1.04 | 0.99 |
| 12 | sarima-untrained_ensemble | 1987.8 | 2552.5 | 0.51 | 0.81 | 1.06 | 1.01 |
| 13 | sarima_s7-noTransform | 2316.8 | 3042.4 | 0.41 | 0.80 | 1.24 | 1.21 |
| 14 | USC-SI_kJalpha | 2635.9 | 3590.1 | 0.36 | 0.77 | 1.41 | 1.43 |

**Table 6:** Summary of best subset models passed on from the validation phase to the testing phase, including explanation of model choice if not strictly based on lowest WIS. Note that THieF_ensemble-train3 was chosen based on cross-validation selection rule of the simplest and not substantially worse model.

| Criteria | Geo Scale | Model |
| --- | --- | --- |
| Best original THieF | states | THieF_6wk-4root |
|  | US | THieF_6wk-noTransform |
| Best (weighted) THieF ensemble | both | THieF_ensemble-train3* |
| Best ARIMA/SARIMA | states | sarima_s7-noTransform |
|  | US | arima_s1-noTransform |
| ARIMA/SARIMA ensemble | NA | sarima_unweighted-mean |
| Untrained THieF ensemble | NA | THieF_ensemble-mean |
| Best original THieF during last 17 weeks | both | THieF_12wk-4root |

**Table 7:** Summary of model performance during the last 17 weeks of the validation phase for averaged states, ordered by ascending WIS.

|  | Model | WIS | MAE | Cov50 | Cov95 | rWIS | rMAE |
| --- | --- | --- | --- | --- | --- | --- | --- |
| 1 | THieF_ensemble-train6.5 | 32.66 | 49.71 | 0.52 | 0.92 | 0.76 | 0.77 |
| 2 | THieF_ensemble-train3 | 32.68 | 49.55 | 0.52 | 0.92 | 0.76 | 0.77 |
| 3 | THieF_ensemble-train10 | 32.70 | 49.87 | 0.51 | 0.92 | 0.76 | 0.77 |
| 4 | THieF_ensemble-mean | 32.71 | 49.19 | 0.53 | 0.92 | 0.76 | 0.76 |
| 5 | THieF_ensemble-train1 | 32.73 | 49.46 | 0.52 | 0.92 | 0.76 | 0.77 |
| 6 | THieF_ensemble-train15 | 32.83 | 50.11 | 0.51 | 0.92 | 0.77 | 0.78 |
| 7 | THieF_ensemble-train20 | 32.99 | 50.34 | 0.50 | 0.92 | 0.77 | 0.78 |
| 8 | THieF_ensemble-train25 | 33.15 | 50.56 | 0.50 | 0.91 | 0.78 | 0.78 |
| 9 | THieF_6wk-4root | 33.36 | 49.31 | 0.49 | 0.91 | 0.78 | 0.77 |
| 10 | THieF_2wk-4root | 34.17 | 51.41 | 0.49 | 0.90 | 0.80 | 0.80 |
| 11 | THieF_2wk-noTransform | 34.36 | 50.18 | 0.51 | 0.88 | 0.80 | 0.78 |
| 12 | THieF_6wk-noTransform | 34.72 | 51.21 | 0.51 | 0.88 | 0.81 | 0.80 |
| 13 | THieF_12wk-4root | 34.90 | 50.27 | 0.49 | 0.90 | 0.82 | 0.78 |
| 14 | THieF_8wk-4root | 35.05 | 51.49 | 0.48 | 0.90 | 0.82 | 0.80 |
| 15 | THieF_1wk-4root | 35.12 | 53.47 | 0.47 | 0.90 | 0.82 | 0.83 |
| 16 | THieF_8wk-noTransform | 35.33 | 52.16 | 0.50 | 0.88 | 0.83 | 0.81 |
| 17 | THieF_3wk-4root | 36.40 | 54.58 | 0.48 | 0.89 | 0.85 | 0.85 |
| 18 | THieF_3wk-noTransform | 36.98 | 53.94 | 0.49 | 0.87 | 0.86 | 0.84 |
| 19 | THieF_4wk-4root | 36.99 | 54.77 | 0.47 | 0.88 | 0.86 | 0.85 |
| 20 | THieF_1wk-noTransform | 37.22 | 54.87 | 0.48 | 0.87 | 0.87 | 0.85 |
| 21 | THieF_4wk-noTransform | 38.18 | 56.00 | 0.48 | 0.86 | 0.89 | 0.87 |
| 22 | THieF_12wk-noTransform | 38.29 | 53.26 | 0.50 | 0.86 | 0.90 | 0.83 |
| 23 | arima_s1-4root | 41.74 | 62.34 | 0.43 | 0.87 | 0.98 | 0.97 |
| 24 | COVIDhub-baseline | 42.78 | 64.41 | 0.72 | 0.97 | 1.00 | 1.00 |
| 25 | sarima_s7-4root | 43.51 | 64.23 | 0.44 | 0.87 | 1.02 | 1.00 |
| 26 | sarima_s7-noTransform | 43.52 | 62.04 | 0.46 | 0.84 | 1.02 | 0.96 |
| 27 | sarima-untrained_ensemble | 45.99 | 67.71 | 0.42 | 0.85 | 1.07 | 1.05 |
| 28 | arima_s1-noTransform | 46.12 | 66.36 | 0.43 | 0.84 | 1.08 | 1.03 |

**Table 8:** Summary of model performance during the last 17 weeks of the validation phase for U.S. national, ordered by ascending WIS.

|  | Model | WIS | MAE | Cov50 | Cov95 | rWIS | rMAE |
| --- | --- | --- | --- | --- | --- | --- | --- |
| 1 | THieF_12wk-4root | 1134.18 | 1632.16 | 0.58 | 0.84 | 0.73 | 0.63 |
| 2 | THieF_ensemble-mean | 1168.76 | 1887.26 | 0.36 | 0.90 | 0.75 | 0.72 |
| 3 | THieF_ensemble-train1 | 1191.03 | 1932.48 | 0.36 | 0.90 | 0.76 | 0.74 |
| 4 | THieF_6wk-4root | 1198.91 | 1799.31 | 0.47 | 0.86 | 0.77 | 0.69 |
| 5 | THieF_ensemble-train3 | 1210.11 | 1970.10 | 0.34 | 0.91 | 0.78 | 0.76 |
| 6 | THieF_6wk-noTransform | 1216.03 | 1906.17 | 0.34 | 0.86 | 0.78 | 0.73 |
| 7 | THieF_ensemble-train6.5 | 1249.32 | 2033.24 | 0.33 | 0.92 | 0.80 | 0.78 |
| 8 | THieF_8wk-noTransform | 1283.13 | 2045.71 | 0.36 | 0.86 | 0.82 | 0.78 |
| 9 | THieF_ensemble-train10 | 1293.18 | 2097.30 | 0.32 | 0.92 | 0.83 | 0.80 |
| 10 | THieF_ensemble-train15 | 1356.59 | 2195.93 | 0.31 | 0.91 | 0.87 | 0.84 |
| 11 | THieF_12wk-noTransform | 1376.62 | 1953.69 | 0.54 | 0.78 | 0.88 | 0.75 |
| 12 | THieF_1wk-noTransform | 1409.47 | 2240.28 | 0.27 | 0.80 | 0.91 | 0.86 |
| 13 | THieF_ensemble-train20 | 1411.90 | 2283.53 | 0.31 | 0.90 | 0.91 | 0.88 |
| 14 | THieF_8wk-4root | 1425.91 | 2111.21 | 0.33 | 0.78 | 0.92 | 0.81 |
| 15 | THieF_4wk-noTransform | 1450.73 | 2141.69 | 0.34 | 0.67 | 0.93 | 0.82 |
| 16 | THieF_ensemble-train25 | 1455.31 | 2349.57 | 0.30 | 0.89 | 0.94 | 0.90 |
| 17 | THieF_2wk-4root | 1456.70 | 2291.52 | 0.26 | 0.83 | 0.94 | 0.88 |
| 18 | THieF_2wk-noTransform | 1457.82 | 2180.36 | 0.31 | 0.73 | 0.94 | 0.84 |
| 19 | THieF_3wk-noTransform | 1487.45 | 2163.36 | 0.32 | 0.66 | 0.96 | 0.83 |
| 20 | THieF_3wk-4root | 1532.98 | 2428.06 | 0.34 | 0.84 | 0.98 | 0.93 |
| 21 | COVIDhub-baseline | 1555.91 | 2606.50 | 0.37 | 0.97 | 1.00 | 1.00 |
| 22 | THieF_1wk-4root | 1565.38 | 2542.24 | 0.27 | 0.85 | 1.01 | 0.97 |
| 23 | THieF_4wk-4root | 1614.42 | 2433.46 | 0.30 | 0.71 | 1.04 | 0.93 |
| 24 | arima_s1-noTransform | 1774.31 | 2639.19 | 0.22 | 0.71 | 1.14 | 1.01 |
| 25 | sarima-untrained_ensemble | 1790.17 | 2855.75 | 0.06 | 0.65 | 1.15 | 1.10 |
| 26 | arima_s1-4root | 1803.20 | 2614.44 | 0.22 | 0.75 | 1.16 | 1.00 |
| 27 | sarima_s7-noTransform | 1922.00 | 2884.58 | 0.15 | 0.65 | 1.24 | 1.11 |
| 28 | sarima_s7-4root | 2155.20 | 3344.10 | 0.17 | 0.73 | 1.38 | 1.28 |

**Table 9:** Summary of model performance during the testing phase stratified by horizon week for averaged states, ordered by ascending WIS.

| Model | Hzn | WIS | MAE | Cov50 | Cov95 | rWIS | rMAE |
| --- | --- | --- | --- | --- | --- | --- | --- |
| COVIDhub-4_week_ensemble | 1 | 16.70 | 25.11 | 0.65 | 0.96 | 0.70 | 0.79 |
| THieF_ensemble-train3 | 1 | 20.32 | 30.04 | 0.55 | 0.93 | 0.85 | 0.94 |
| THieF_ensemble-mean | 1 | 20.76 | 30.52 | 0.56 | 0.93 | 0.87 | 0.96 |
| CU-select | 1 | 20.81 | 27.91 | 0.33 | 0.74 | 0.87 | 0.88 |
| THieF_6wk-4root | 1 | 22.50 | 32.97 | 0.49 | 0.89 | 0.94 | 1.03 |
| GT-DeepCOVID | 1 | 22.65 | 30.64 | 0.30 | 0.63 | 0.95 | 0.96 |
| sarima_s7-noTransform | 1 | 23.11 | 31.92 | 0.57 | 0.89 | 0.97 | 1.00 |
| sarima-untrained_ensemble | 1 | 23.12 | 31.75 | 0.60 | 0.89 | 0.97 | 1.00 |
| JHUAPL-Bucky | 1 | 23.38 | 34.92 | 0.46 | 0.89 | 0.98 | 1.10 |
| COVIDhub-baseline | 1 | 23.92 | 31.87 | 0.80 | 0.97 | 1.00 | 1.00 |
| arima_s1-noTransform | 1 | 23.98 | 32.81 | 0.60 | 0.89 | 1.00 | 1.03 |
| THieF_12wk-4root | 1 | 24.28 | 35.21 | 0.48 | 0.89 | 1.02 | 1.10 |
| THieF_6wk-noTransform | 1 | 25.17 | 36.41 | 0.53 | 0.89 | 1.05 | 1.14 |
| USC-SI_kJalpha | 1 | 29.63 | 37.54 | 0.22 | 0.55 | 1.24 | 1.18 |
| COVIDhub-4_week_ensemble | 2 | 25.42 | 37.90 | 0.66 | 0.95 | 0.69 | 0.77 |
| THieF_6wk-4root | 2 | 30.10 | 43.94 | 0.52 | 0.91 | 0.81 | 0.89 |
| THieF_ensemble-train3 | 2 | 30.66 | 44.49 | 0.57 | 0.91 | 0.83 | 0.90 |
| THieF_ensemble-mean | 2 | 31.04 | 44.80 | 0.58 | 0.91 | 0.84 | 0.91 |
| CU-select | 2 | 31.20 | 43.54 | 0.34 | 0.76 | 0.84 | 0.88 |
| THieF_12wk-4root | 2 | 31.29 | 45.58 | 0.51 | 0.90 | 0.85 | 0.92 |
| THieF_6wk-noTransform | 2 | 35.00 | 50.27 | 0.56 | 0.88 | 0.95 | 1.02 |
| JHUAPL-Bucky | 2 | 35.10 | 52.62 | 0.50 | 0.91 | 0.95 | 1.06 |
| GT-DeepCOVID | 2 | 36.53 | 48.38 | 0.29 | 0.61 | 0.99 | 0.98 |
| sarima-untrained_ensemble | 2 | 36.65 | 49.30 | 0.61 | 0.87 | 0.99 | 1.00 |
| COVIDhub-baseline | 2 | 36.95 | 49.48 | 0.80 | 0.96 | 1.00 | 1.00 |
| sarima_s7-noTransform | 2 | 37.32 | 50.28 | 0.60 | 0.87 | 1.01 | 1.02 |
| arima_s1-noTransform | 2 | 37.78 | 50.84 | 0.61 | 0.87 | 1.02 | 1.03 |
| USC-SI_kJalpha | 2 | 50.42 | 63.90 | 0.21 | 0.52 | 1.36 | 1.29 |
| COVIDhub-4_week_ensemble | 3 | 35.22 | 52.52 | 0.64 | 0.94 | 0.73 | 0.79 |
| THieF_6wk-4root | 3 | 38.98 | 56.36 | 0.52 | 0.90 | 0.80 | 0.85 |
| THieF_12wk-4root | 3 | 39.40 | 57.34 | 0.51 | 0.90 | 0.81 | 0.86 |
| CU-select | 3 | 41.03 | 60.37 | 0.32 | 0.75 | 0.85 | 0.91 |
| THieF_ensemble-train3 | 3 | 41.60 | 59.88 | 0.56 | 0.90 | 0.86 | 0.90 |
| THieF_ensemble-mean | 3 | 41.85 | 60.07 | 0.58 | 0.90 | 0.86 | 0.91 |
| THieF_6wk-noTransform | 3 | 45.11 | 64.82 | 0.55 | 0.87 | 0.93 | 0.98 |
| COVIDhub-baseline | 3 | 48.54 | 66.30 | 0.79 | 0.95 | 1.00 | 1.00 |
| sarima-untrained_ensemble | 3 | 49.27 | 66.33 | 0.60 | 0.85 | 1.02 | 1.00 |
| arima_s1-noTransform | 3 | 50.20 | 67.64 | 0.60 | 0.85 | 1.03 | 1.02 |
| sarima_s7-noTransform | 3 | 51.31 | 69.07 | 0.61 | 0.86 | 1.06 | 1.04 |
| JHUAPL-Bucky | 3 | 52.06 | 76.94 | 0.49 | 0.89 | 1.07 | 1.16 |
| GT-DeepCOVID | 3 | 52.73 | 68.51 | 0.26 | 0.57 | 1.09 | 1.03 |
| USC-SI_kJalpha | 3 | 83.34 | 106.44 | 0.20 | 0.47 | 1.72 | 1.61 |
| COVIDhub-4_week_ensemble | 4 | 44.39 | 65.43 | 0.63 | 0.93 | 0.76 | 0.81 |
| THieF_12wk-4root | 4 | 46.84 | 68.49 | 0.50 | 0.89 | 0.80 | 0.84 |
| THieF_6wk-4root | 4 | 47.53 | 68.48 | 0.52 | 0.88 | 0.81 | 0.84 |
| CU-select | 4 | 47.54 | 69.72 | 0.31 | 0.73 | 0.81 | 0.86 |
| THieF_ensemble-train3 | 4 | 51.04 | 73.47 | 0.55 | 0.89 | 0.87 | 0.90 |
| THieF_ensemble-mean | 4 | 51.11 | 73.39 | 0.57 | 0.89 | 0.88 | 0.90 |
| THieF_6wk-noTransform | 4 | 53.80 | 77.46 | 0.55 | 0.86 | 0.92 | 0.95 |
| COVIDhub-baseline | 4 | 58.36 | 81.23 | 0.77 | 0.95 | 1.00 | 1.00 |
| sarima-untrained_ensemble | 4 | 60.56 | 81.92 | 0.59 | 0.84 | 1.04 | 1.01 |
| arima_s1-noTransform | 4 | 61.05 | 82.67 | 0.58 | 0.83 | 1.05 | 1.02 |
| sarima_s7-noTransform | 4 | 63.89 | 86.44 | 0.61 | 0.85 | 1.09 | 1.06 |
| GT-DeepCOVID | 4 | 66.48 | 85.93 | 0.25 | 0.55 | 1.14 | 1.06 |
| JHUAPL-Bucky | 4 | 68.14 | 98.29 | 0.47 | 0.88 | 1.17 | 1.21 |
| USC-SI_kJalpha | 4 | 122.37 | 157.37 | 0.20 | 0.45 | 2.10 | 1.94 |

**Table 10:**
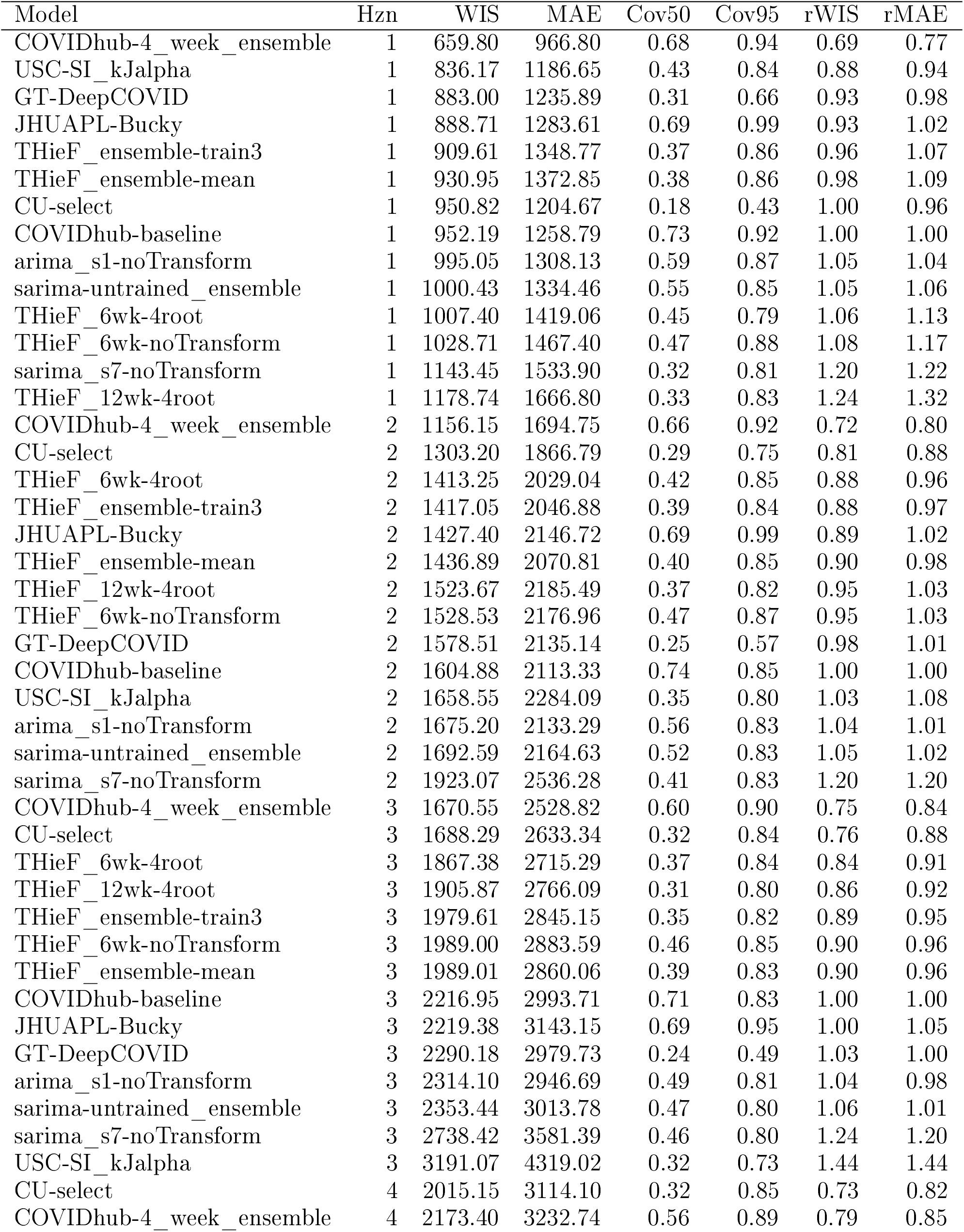
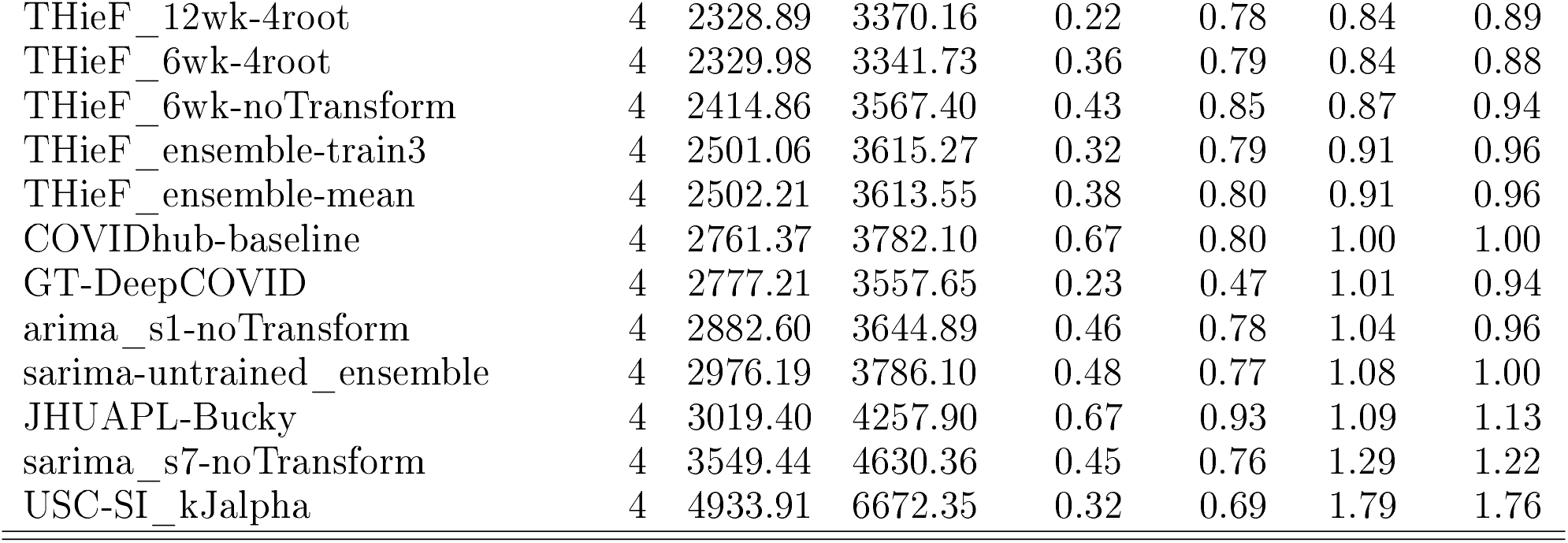
Summary of model performance during the testing phase stratified by horizon week for U.S. national, ordered by ascending WIS.

| Model | Hzn | WIS | MAE | Cov50 | Cov95 | rWIS | rMAE |
| --- | --- | --- | --- | --- | --- | --- | --- |
| COVIDhub-4_week_ensemble | 1 | 659.80 | 966.80 | 0.68 | 0.94 | 0.69 | 0.77 |
| USC-SI_kJalpha | 1 | 836.17 | 1186.65 | 0.43 | 0.84 | 0.88 | 0.94 |
| GT-DeepCOVID | 1 | 883.00 | 1235.89 | 0.31 | 0.66 | 0.93 | 0.98 |
| JHUAPL-Bucky | 1 | 888.71 | 1283.61 | 0.69 | 0.99 | 0.93 | 1.02 |
| THieF_ensemble-train3 | 1 | 909.61 | 1348.77 | 0.37 | 0.86 | 0.96 | 1.07 |
| THieF_ensemble-mean | 1 | 930.95 | 1372.85 | 0.38 | 0.86 | 0.98 | 1.09 |
| CU-select | 1 | 950.82 | 1204.67 | 0.18 | 0.43 | 1.00 | 0.96 |
| COVIDhub-baseline | 1 | 952.19 | 1258.79 | 0.73 | 0.92 | 1.00 | 1.00 |
| arima_s1-noTransform | 1 | 995.05 | 1308.13 | 0.59 | 0.87 | 1.05 | 1.04 |
| sarima-untrained_ensemble | 1 | 1000.43 | 1334.46 | 0.55 | 0.85 | 1.05 | 1.06 |
| THieF_6wk-4root | 1 | 1007.40 | 1419.06 | 0.45 | 0.79 | 1.06 | 1.13 |
| THieF_6wk-noTransform | 1 | 1028.71 | 1467.40 | 0.47 | 0.88 | 1.08 | 1.17 |
| sarima_s7-noTransform | 1 | 1143.45 | 1533.90 | 0.32 | 0.81 | 1.20 | 1.22 |
| THieF_12wk-4root | 1 | 1178.74 | 1666.80 | 0.33 | 0.83 | 1.24 | 1.32 |
| COVIDhub-4_week_ensemble | 2 | 1156.15 | 1694.75 | 0.66 | 0.92 | 0.72 | 0.80 |
| CU-select | 2 | 1303.20 | 1866.79 | 0.29 | 0.75 | 0.81 | 0.88 |
| THieF_6wk-4root | 2 | 1413.25 | 2029.04 | 0.42 | 0.85 | 0.88 | 0.96 |
| THieF_ensemble-train3 | 2 | 1417.05 | 2046.88 | 0.39 | 0.84 | 0.88 | 0.97 |
| JHUAPL-Bucky | 2 | 1427.40 | 2146.72 | 0.69 | 0.99 | 0.89 | 1.02 |
| THieF_ensemble-mean | 2 | 1436.89 | 2070.81 | 0.40 | 0.85 | 0.90 | 0.98 |
| THieF_12wk-4root | 2 | 1523.67 | 2185.49 | 0.37 | 0.82 | 0.95 | 1.03 |
| THieF_6wk-noTransform | 2 | 1528.53 | 2176.96 | 0.47 | 0.87 | 0.95 | 1.03 |
| GT-DeepCOVID | 2 | 1578.51 | 2135.14 | 0.25 | 0.57 | 0.98 | 1.01 |
| COVIDhub-baseline | 2 | 1604.88 | 2113.33 | 0.74 | 0.85 | 1.00 | 1.00 |
| USC-SI_kJalpha | 2 | 1658.55 | 2284.09 | 0.35 | 0.80 | 1.03 | 1.08 |
| arima_s1-noTransform | 2 | 1675.20 | 2133.29 | 0.56 | 0.83 | 1.04 | 1.01 |
| sarima-untrained_ensemble | 2 | 1692.59 | 2164.63 | 0.52 | 0.83 | 1.05 | 1.02 |
| sarima_s7-noTransform | 2 | 1923.07 | 2536.28 | 0.41 | 0.83 | 1.20 | 1.20 |
| COVIDhub-4_week_ensemble | 3 | 1670.55 | 2528.82 | 0.60 | 0.90 | 0.75 | 0.84 |
| CU-select | 3 | 1688.29 | 2633.34 | 0.32 | 0.84 | 0.76 | 0.88 |
| THieF_6wk-4root | 3 | 1867.38 | 2715.29 | 0.37 | 0.84 | 0.84 | 0.91 |
| THieF_12wk-4root | 3 | 1905.87 | 2766.09 | 0.31 | 0.80 | 0.86 | 0.92 |
| THieF_ensemble-train3 | 3 | 1979.61 | 2845.15 | 0.35 | 0.82 | 0.89 | 0.95 |
| THieF_6wk-noTransform | 3 | 1989.00 | 2883.59 | 0.46 | 0.85 | 0.90 | 0.96 |
| THieF_ensemble-mean | 3 | 1989.01 | 2860.06 | 0.39 | 0.83 | 0.90 | 0.96 |
| COVIDhub-baseline | 3 | 2216.95 | 2993.71 | 0.71 | 0.83 | 1.00 | 1.00 |
| JHUAPL-Bucky | 3 | 2219.38 | 3143.15 | 0.69 | 0.95 | 1.00 | 1.05 |
| GT-DeepCOVID | 3 | 2290.18 | 2979.73 | 0.24 | 0.49 | 1.03 | 1.00 |
| arima_s1-noTransform | 3 | 2314.10 | 2946.69 | 0.49 | 0.81 | 1.04 | 0.98 |
| sarima-untrained_ensemble | 3 | 2353.44 | 3013.78 | 0.47 | 0.80 | 1.06 | 1.01 |
| sarima_s7-noTransform | 3 | 2738.42 | 3581.39 | 0.46 | 0.80 | 1.24 | 1.20 |
| USC-SI_kJalpha | 3 | 3191.07 | 4319.02 | 0.32 | 0.73 | 1.44 | 1.44 |
| CU-select | 4 | 2015.15 | 3114.10 | 0.32 | 0.85 | 0.73 | 0.82 |
| COVIDhub-4_week_ensemble | 4 | 2173.40 | 3232.74 | 0.56 | 0.89 | 0.79 | 0.85 |
| THieF_12wk-4root | 4 | 2328.89 | 3370.16 | 0.22 | 0.78 | 0.84 | 0.89 |
| THieF_6wk-4root | 4 | 2329.98 | 3341.73 | 0.36 | 0.79 | 0.84 | 0.88 |
| THieF_6wk-noTransform | 4 | 2414.86 | 3567.40 | 0.43 | 0.85 | 0.87 | 0.94 |
| THieF_ensemble-train3 | 4 | 2501.06 | 3615.27 | 0.32 | 0.79 | 0.91 | 0.96 |
| THieF_ensemble-mean | 4 | 2502.21 | 3613.55 | 0.38 | 0.80 | 0.91 | 0.96 |
| COVIDhub-baseline | 4 | 2761.37 | 3782.10 | 0.67 | 0.80 | 1.00 | 1.00 |
| GT-DeepCOVID | 4 | 2777.21 | 3557.65 | 0.23 | 0.47 | 1.01 | 0.94 |
| arima_s1-noTransform | 4 | 2882.60 | 3644.89 | 0.46 | 0.78 | 1.04 | 0.96 |
| sarima-untrained_ensemble | 4 | 2976.19 | 3786.10 | 0.48 | 0.77 | 1.08 | 1.00 |
| JHUAPL-Bucky | 4 | 3019.40 | 4257.90 | 0.67 | 0.93 | 1.09 | 1.13 |
| sarima_s7-noTransform | 4 | 3549.44 | 4630.36 | 0.45 | 0.76 | 1.29 | 1.22 |
| USC-SI_kJalpha | 4 | 4933.91 | 6672.35 | 0.32 | 0.69 | 1.79 | 1.76 |

**Table 11:**
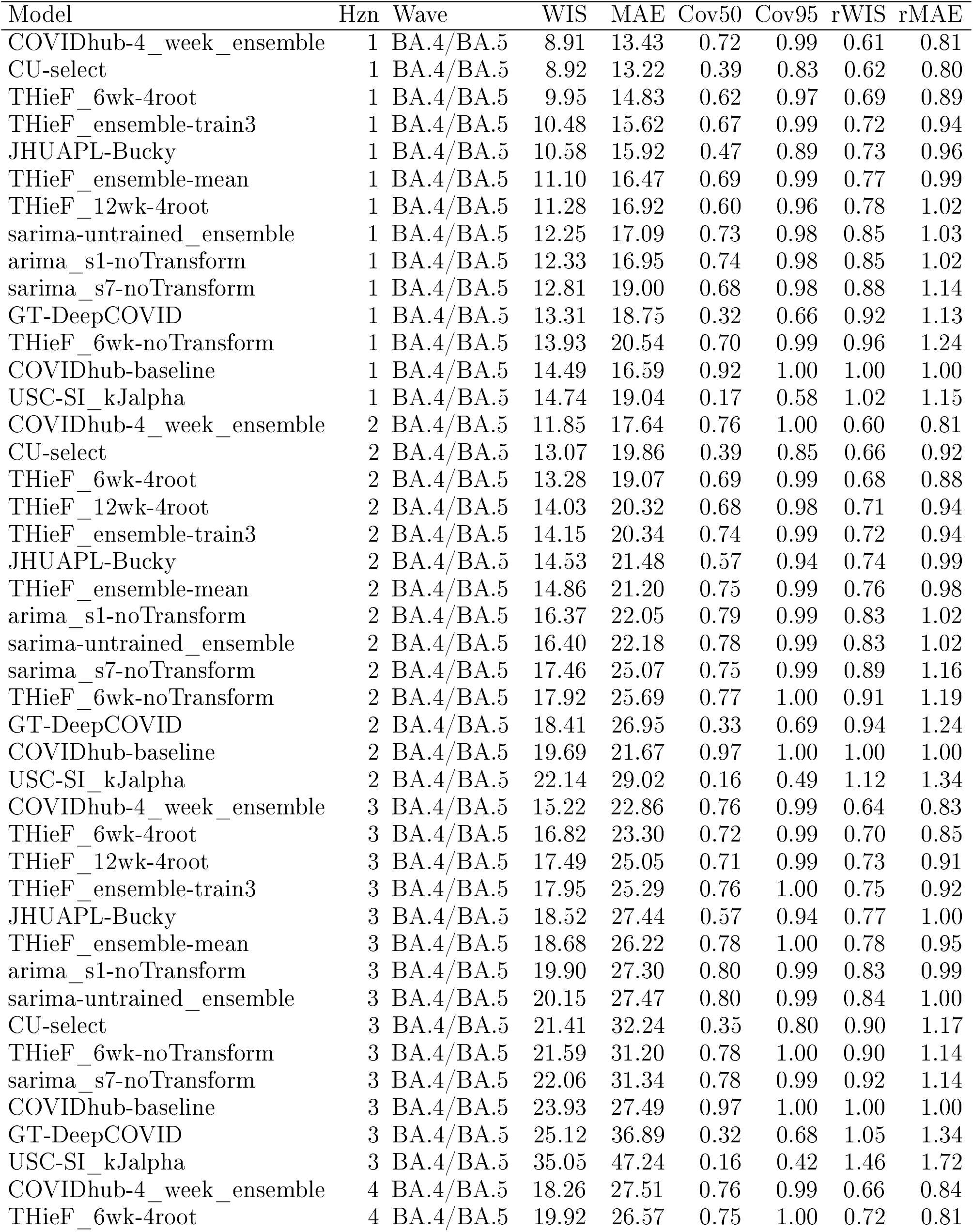
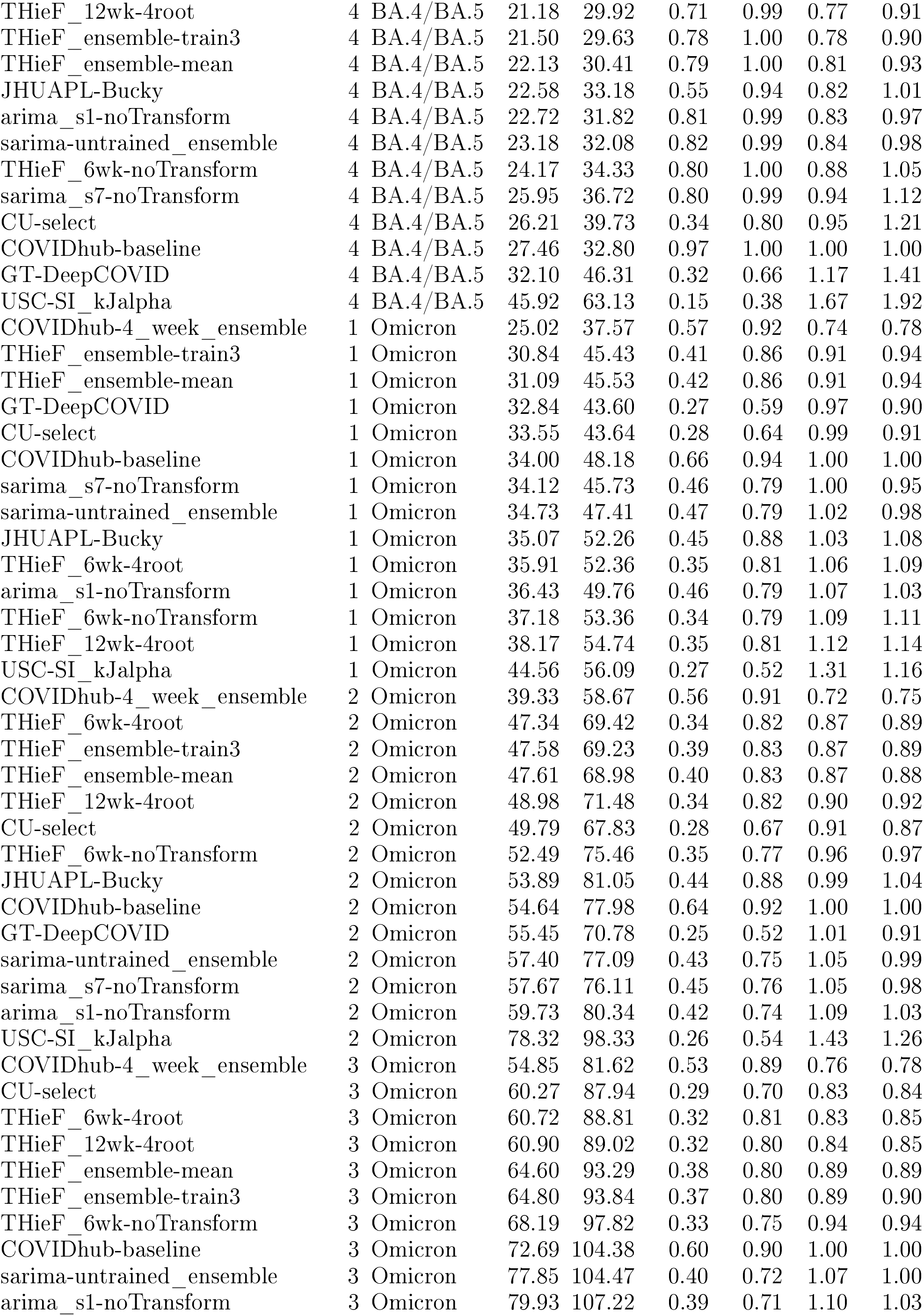
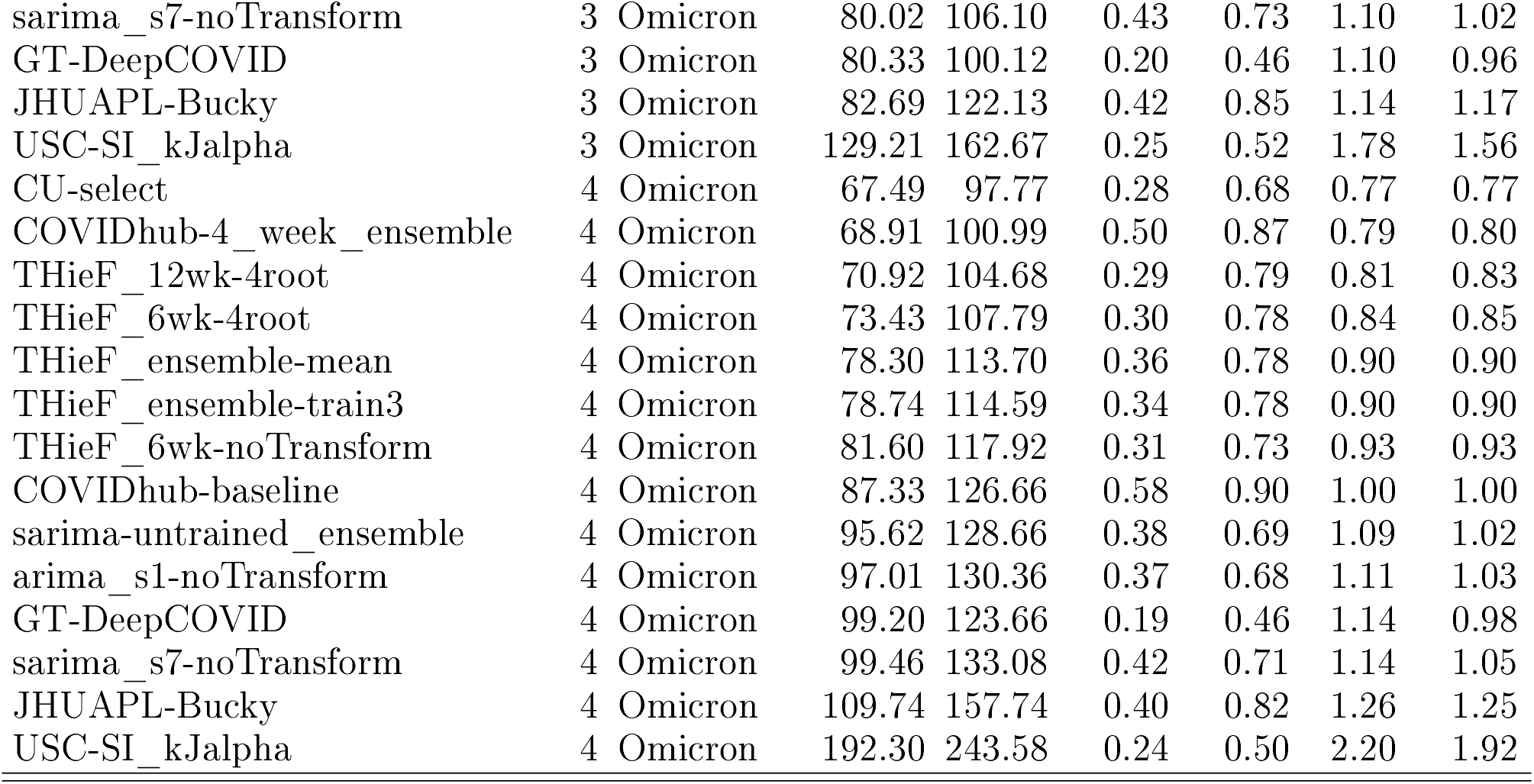
Summary of model performance during the testing phase stratified by pandemic wave and horizon week for averaged states, ordered by ascending WIS.

**Table 12:**
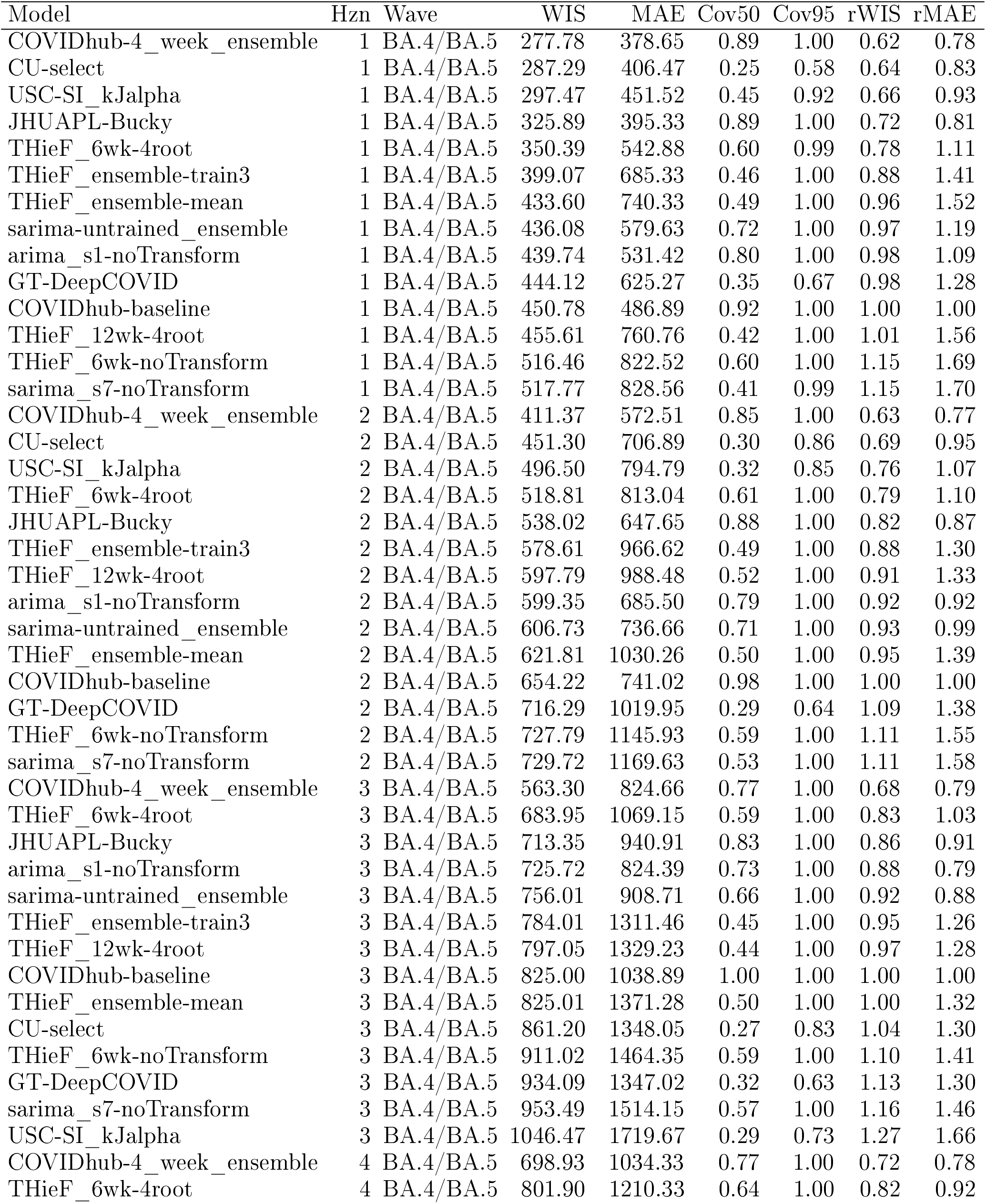
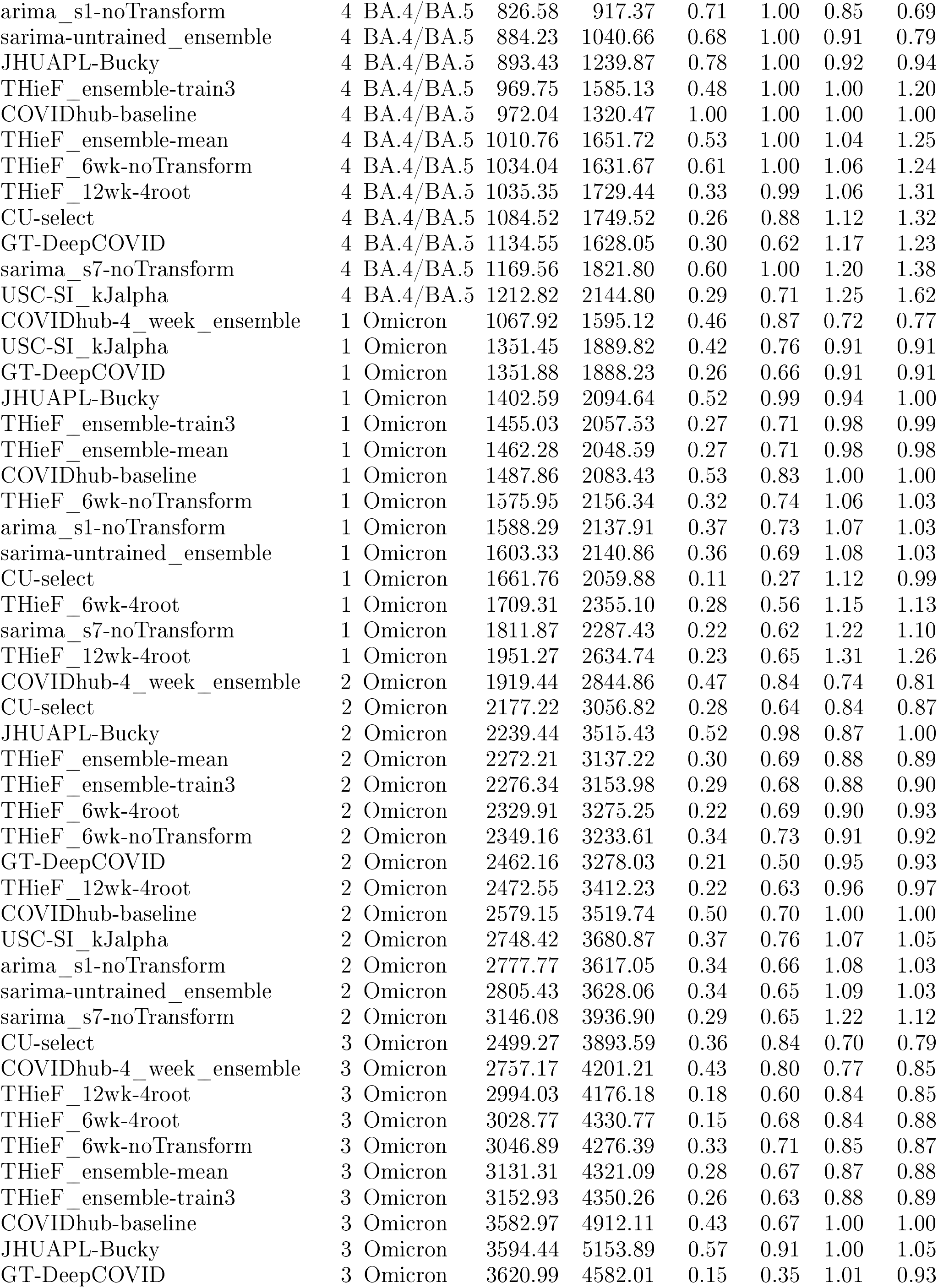
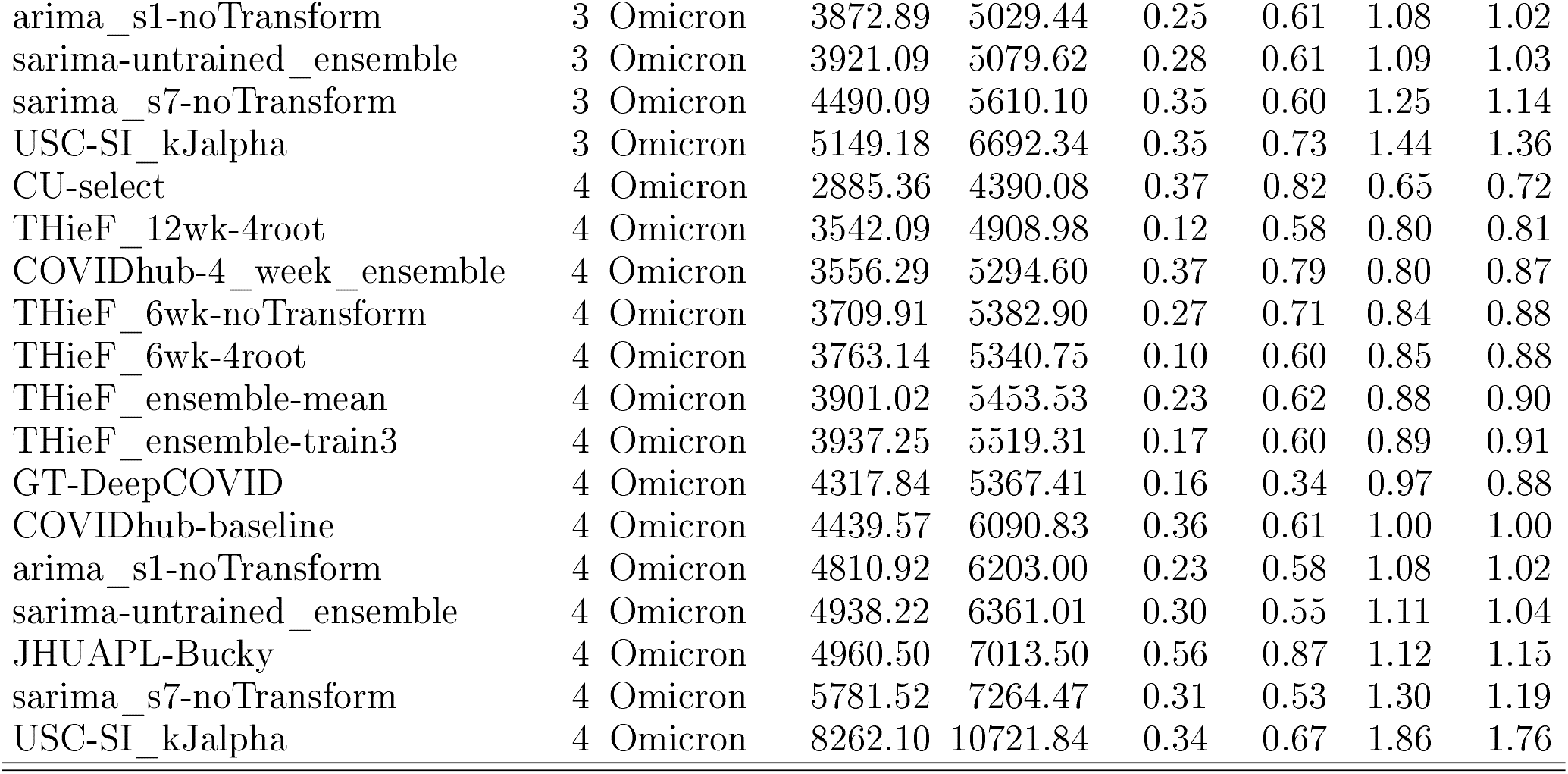
Summary of model performance during the testing phase stratified by pandemic wave and horizon week for U.S. national, ordered by ascending WIS.

#### 3.2.2 Model performance by horizon week

The by-horizon evaluation showed similar model rankings to those of the overall evaluation (Figure 4 and Supplemental Tables 9 and 10). The COVIDhub-4_week_ensemble had lower WIS values for most horizons and geographic scales compared to the other thirteen models while THieF_6wk-4root or CU-select (for states or US national, respectively) tended to have the second-lowest WIS for most horizons, beating the corresponding THieF_6wk-noTransform model at every horizon. Additionally, for all but the 1-week ahead horizon, the top seven models beat the COVIDhub-baseline in terms of lowest WIS for both geographic scales. However, aggregating the forecasts by horizon illustrates some important differences in accuracy between horizon weeks.

**Figure 4:**
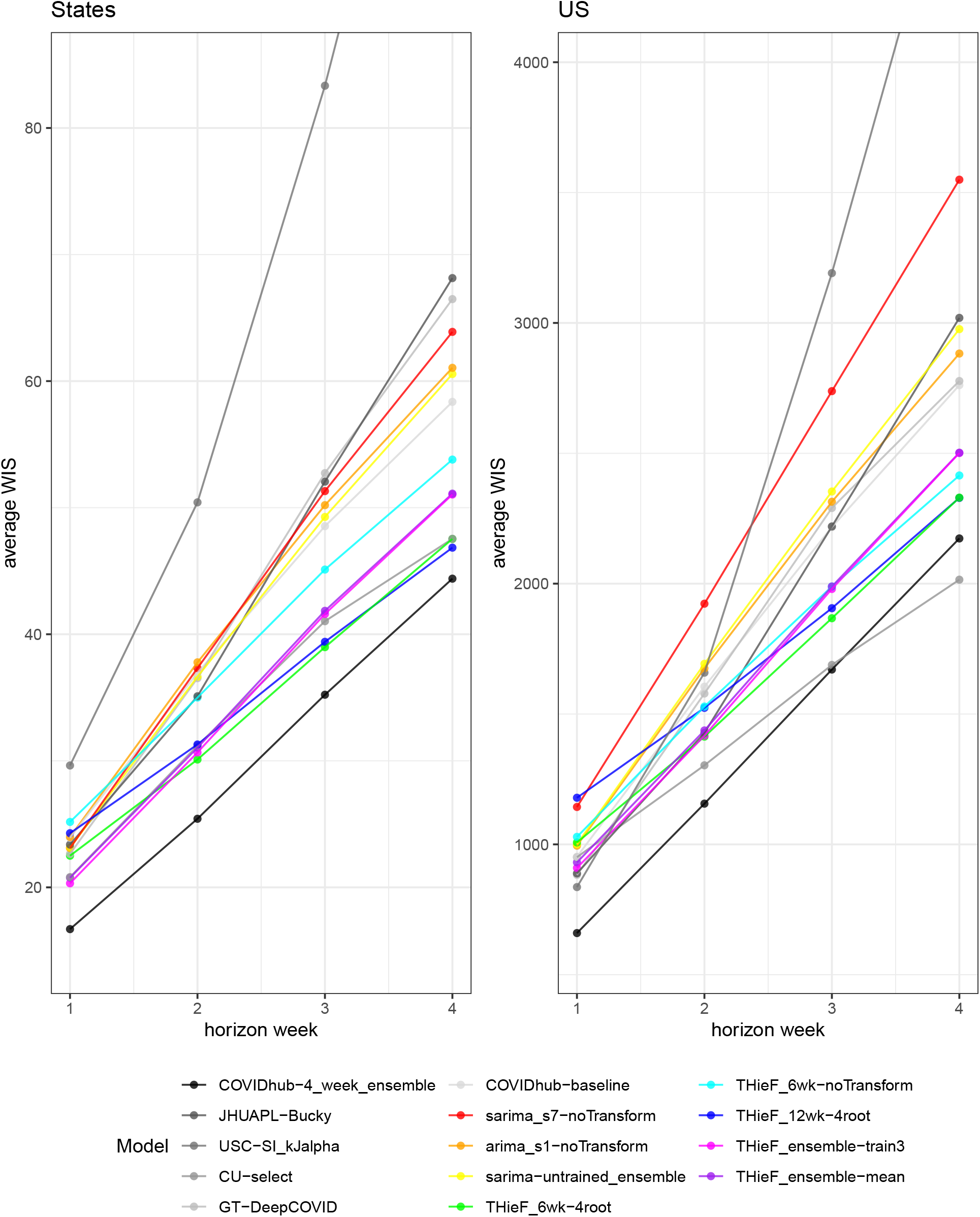
Average WIS plotted by horizon week during the testing phase for each model at both geographic scales

For both geographic scales, the THieF ensembles outperformed CU-select and the fourth root, original THieF models at shorter horizons while the reverse is true at longer ones Figure 4). Notably, THieF_12wk-4root had one of the highest WIS values at the 1-week horizon but had the second lowest WIS at the 4-week horizon. CU-select also performed as well or better than the COVIDhub-4_week_ensemble for the 3- and 4-week ahead horizons. In general, there were several models that switched ranking order as horizon week increased, shown by crossing lines in Figure 4. Meanwhile, the difference in coverage rates between the average states and U.S. national scales remained similar across all horizons and models (see Supplemental Tables 9 and 10).

#### 3.2.3 Model performance by pandemic wave

The COVIDhub-4_week_ensemble yet again generally ranked first for lowest WIS and MAE for the by—pandemic wave evaluation, and was better than all other models at every horizon except CU-select for longer horizons during Omicron (see Figure 5 and Supplemental Tables 11 and 12). CU-select s substantially better performance was likely due to its close adherence to the observed hospitalizations even at long horizons during this wave (see Supplemental Forecast Plots). The rankings of the other models by lowest WIS varied for each combination of pandemic wave, geographic scale, and horizon; though CU-select, the fourth root original THieF models, and the THieF ensembles usually ranked in the top half of models while sarima_s7-noTransform, JHUAPL-Bucky, USC-SI_kJalpha, GT-DeepCOVID, and the COVIDhub-baseline usually ranked near the bottom. Model performance tended to be consistent for the same pandemic wave, regardless of geographic scale (Figure 5).

**Figure 5:**
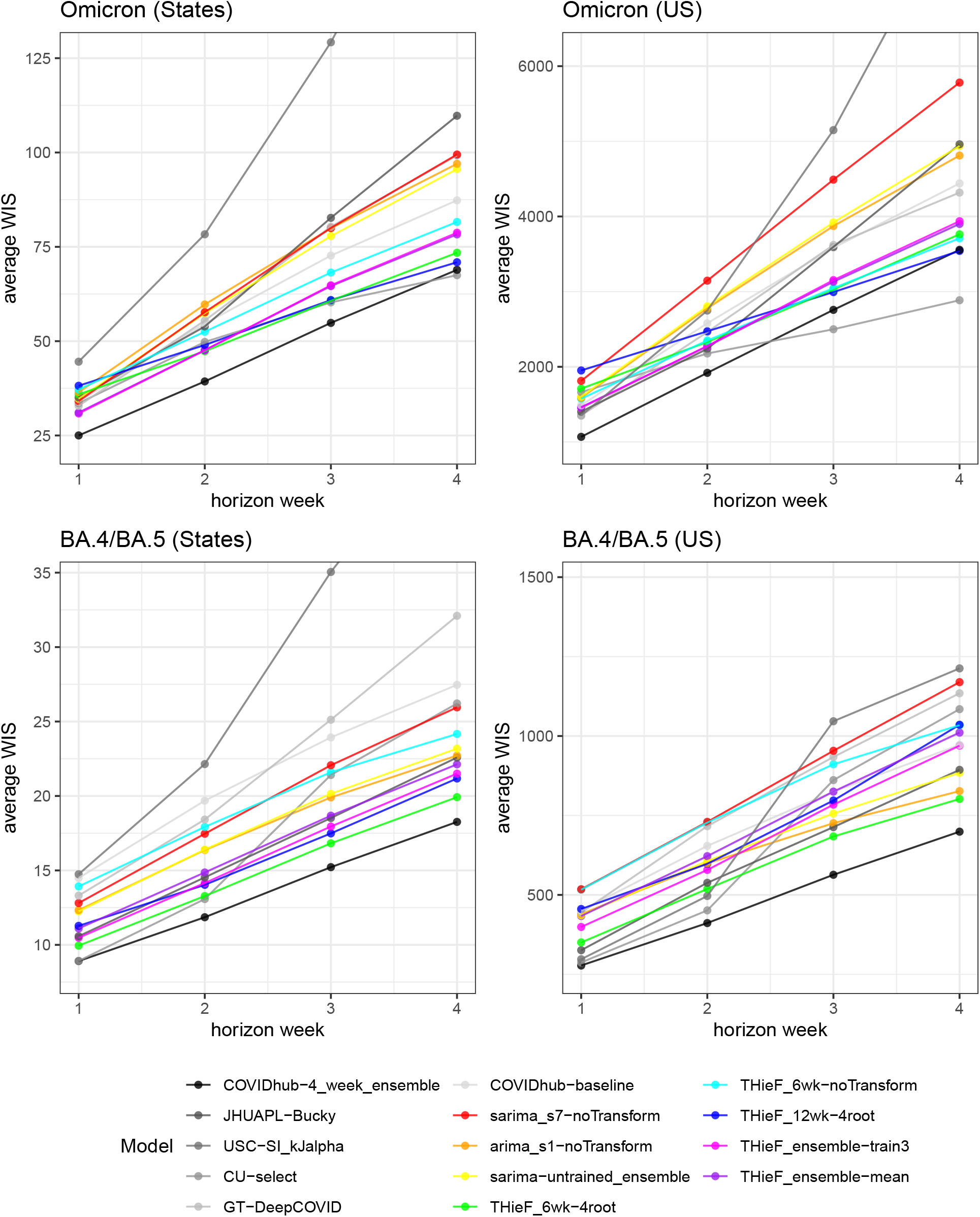
Average WIS plotted by pandemic wave and horizon week during the testing phase for each model at both geographic scales

For the large Omicron wave, we once again observed the THieF ensembles performing better at shorter horizons while CU-select and the fourth root THieF models performed better at longer ones (Figure 5). Similar to the by-horizon evaluation, THieF_12wk-4root had the highest WIS of the fourteen models at the 1-week ahead horizon but shifted to have one of the lowest WIS values at the 4-week horizon; this likely was the result of its longer range forecasts better adhering to the eventually observed COVID-19 hospitalizations than most other models (see Supplemental Forecast Plots). THieF_6wk-noTransform and GT-DeepCOVID also showed better relative performance during this wave compared to BA.4/BA.5, especially for the U.S. national scale. Most models had high WIS values and below-nominal coverage rates during this wave for their respective geographic scale (Figure 5, Supplemental Tables 11 and 12).

For the smaller BA.4/BA.5 variant wave, model rankings by WIS were more distinct and tended to remain static from one horizon week to another, with COVIDhub-4_week_ensemble and THieF_6wk-4root leading the pack (Figure 5). However, CU-select displayed a sharp increase in WIS starting at the 3-week ahead horizon for this wave, causing it to drop from second place to near the bottom of the ranks for the longer horizons. Based on the Supplemental Forecast Plots, CU-select seems to have had difficulty matching trends at longer horizons for times of greater stability compared to the other models. Coverage rates were also high overall for both geographic scales, most models attaining nominal coverage rates or higher (Supplemental Tables 11 and 12). Both of these patterns were likely a result of the wave being fairly stable and small in terms of its magnitude and rate of change.

#### 3.2.4 Model performance by forecast week

Many patterns observed in evaluations described in previous subsection persisted for the by forecast week evaluation. The COVIDhub-4_week_ensemble consistently was one of the the top few, if not the top, models for both WIS and 95% coverage for both geographic scales (Figures 6 and 7). The one exception occurred near the crest of the Omicron wave when this model had either middling or some of the highest WIS values at the 1- and 4-week ahead horizons respectively. During these instances, other well-performing models like CU-select, THieF_6wk-4root, THieF_6wk-noTransform, and THieF_12wk-4root assumed the top spots in the rankings (Figures 6 and 7).

**Figure 6:**
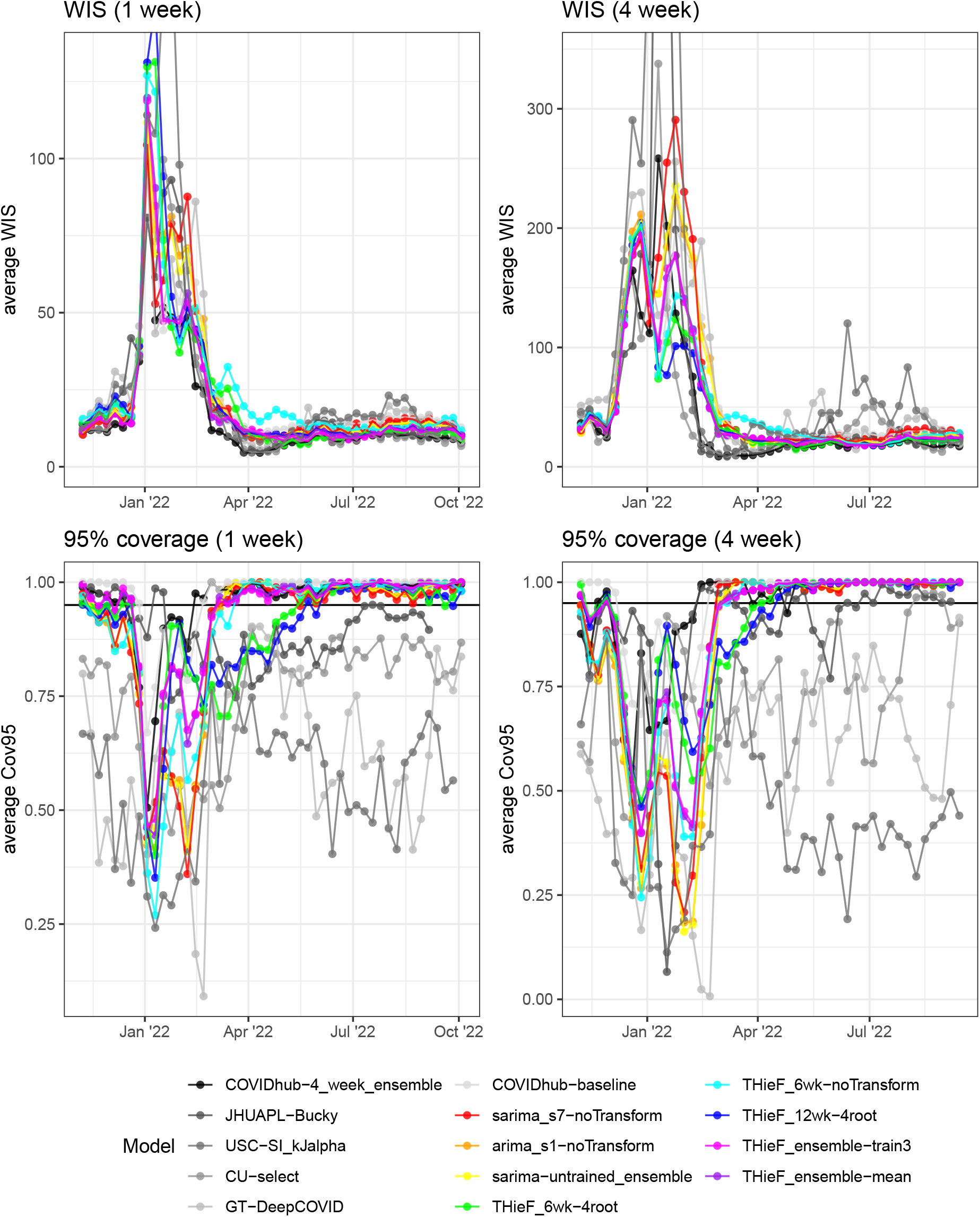
Average h-week ahead WIS and 95% PI coverage plotted by forecast week during the testing phase for each model for averaged states. The plotted WIS values generally follow the shape of the incident hospitalization truth data, with greater values occurring during times of high incidence and rapid change. The 95% coverage rates tend to follow a vertically flipped transformation of the truth data, with below-nominal coverage rates observed during times of high incidence and rapid change.

**Figure 7:**
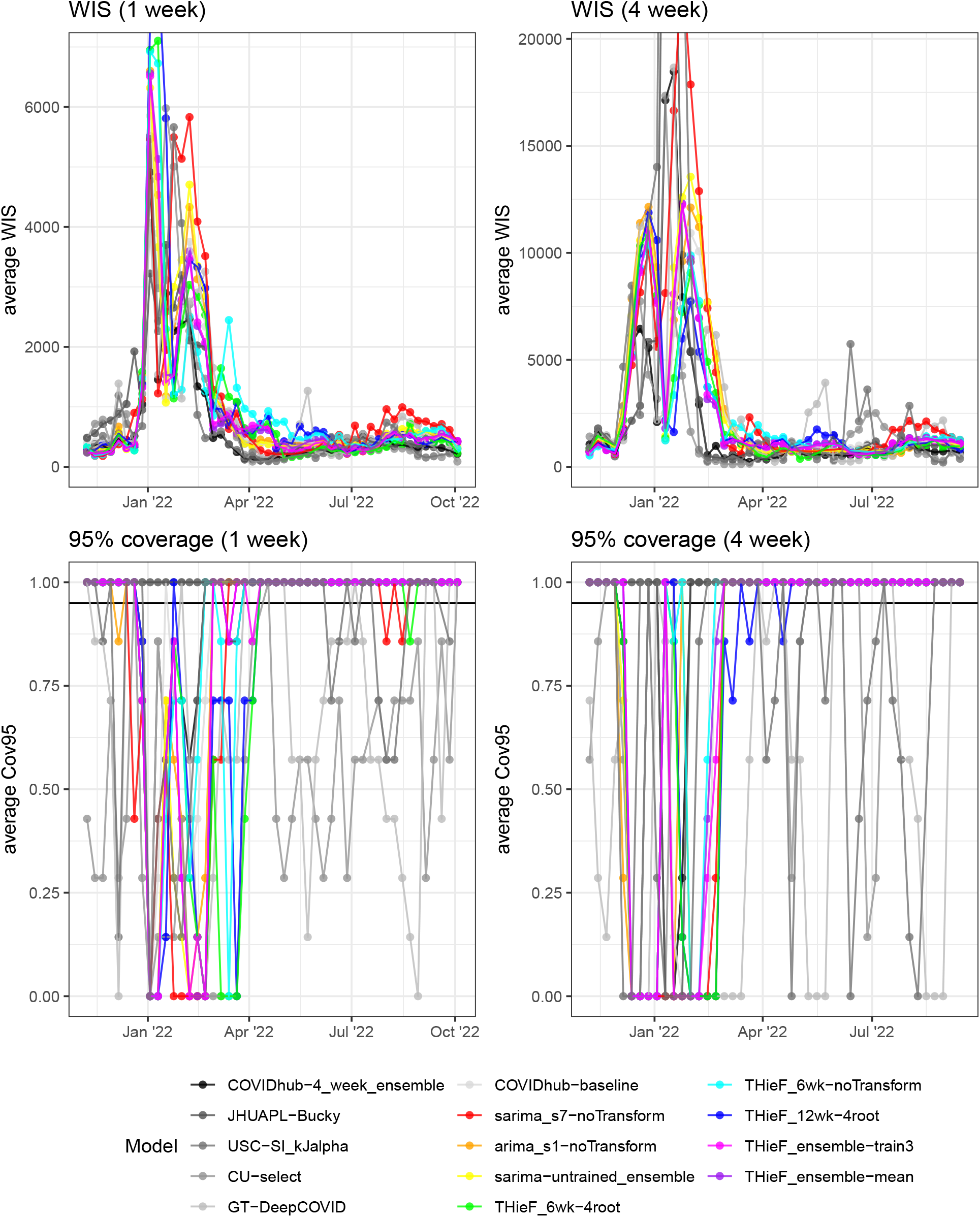
Average h-week ahead WIS and 95% PI coverage plotted by forecast week during the testing phase for each model for the entire U.S.

The two THieF ensembles, which rarely beat the COVIDhub-4_week_ensemble in terms of WIS, demonstrated incredibly stable performance, rarely ranking below seventh. They, like the COVIDhub-baseline, displayed great consistency from week-to-week during times of high incidence and rapid change (Figures 6 and 7). Additionally, model rankings could completely flip at times, with some of the worst-performing models displaying the best WIS and 95% coverage rates during times of rapid change. Generally, though, the observed incident hospitalizations are positively correlated with WIS and negatively correlated with 95% prediction interval coverage (Figures 6 and 7). Further, both CU-select fourth root original THieF models displayed strangely low 95% coverage for averaged states following the decline of the Omicron wave at both plotted horizons, especially since many other models experienced significantly higher coverage rates during this time, while still retaining relatively good WIS values (Figures 6 and 7). Comparisons among models for U.S. national 95% coverage by forecast week were harder to make since this location scale only included seven 95% interval forecasts for one location (compared to seven per 52 locations for averaged states), so we simply allow the plots in Figure 7 to speak for themselves.

#### 3.2.5 Model performance by location

For the by-location evaluation the top seven models had better or equivalent performance compared to the COVIDhub-baseline (Figure 8). The COVIDhub-4_week_ensemble was first place yet again for the evaluation while the other top models demonstrated close WIS values across all locations. CU-select also performed very well, beating the COVIDhub-4_week_ensemble by more than 0.05 relative WIS in over 18% of locations. As before, this good performance was likely owed to its close adherence to the truth data during the Omicron wave. THieF_6wk-4root (third place) was marginally better than the four models ranked below it with lower relative WIS by a difference of more than 0.05 in over 18% of locations. THieF_12wk-4root ranked fourth) had several locations for which it outperformed the COVIDhub-4_week_ensemble, CU-select, THieF_6wk-4root (Figure 8).

**Figure 8:**
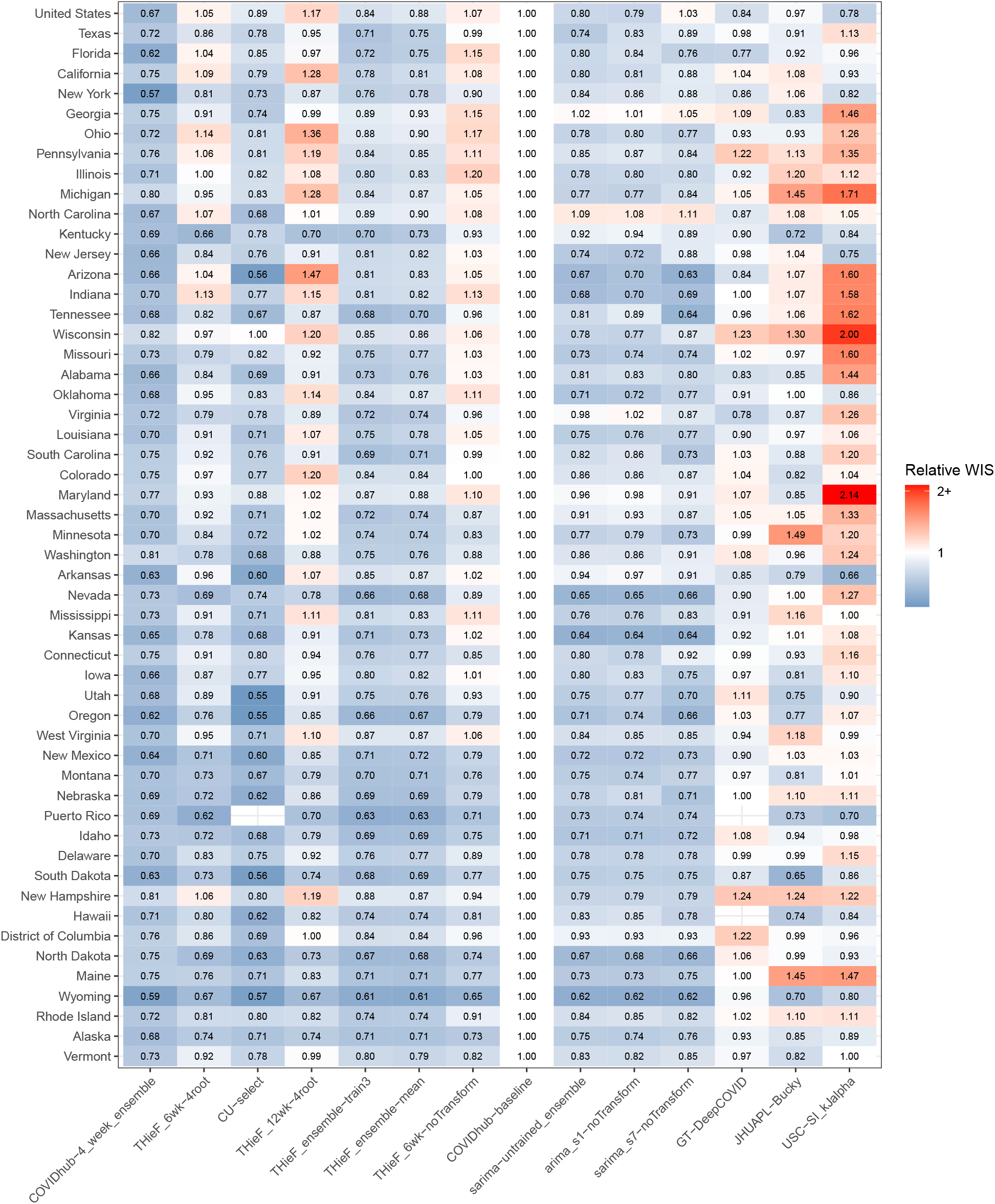
Relative WIS (rWIS) plotted by location during the testing phase for each model across all horizons. Models are ordered by lowest-to-highest rWIS on the x-axis, locations by descending cumulative hospitalizations on the y-axis. Shades of blue indicate better performance than the naive baseline, while red indicates worse performance.

**Figure 9:**
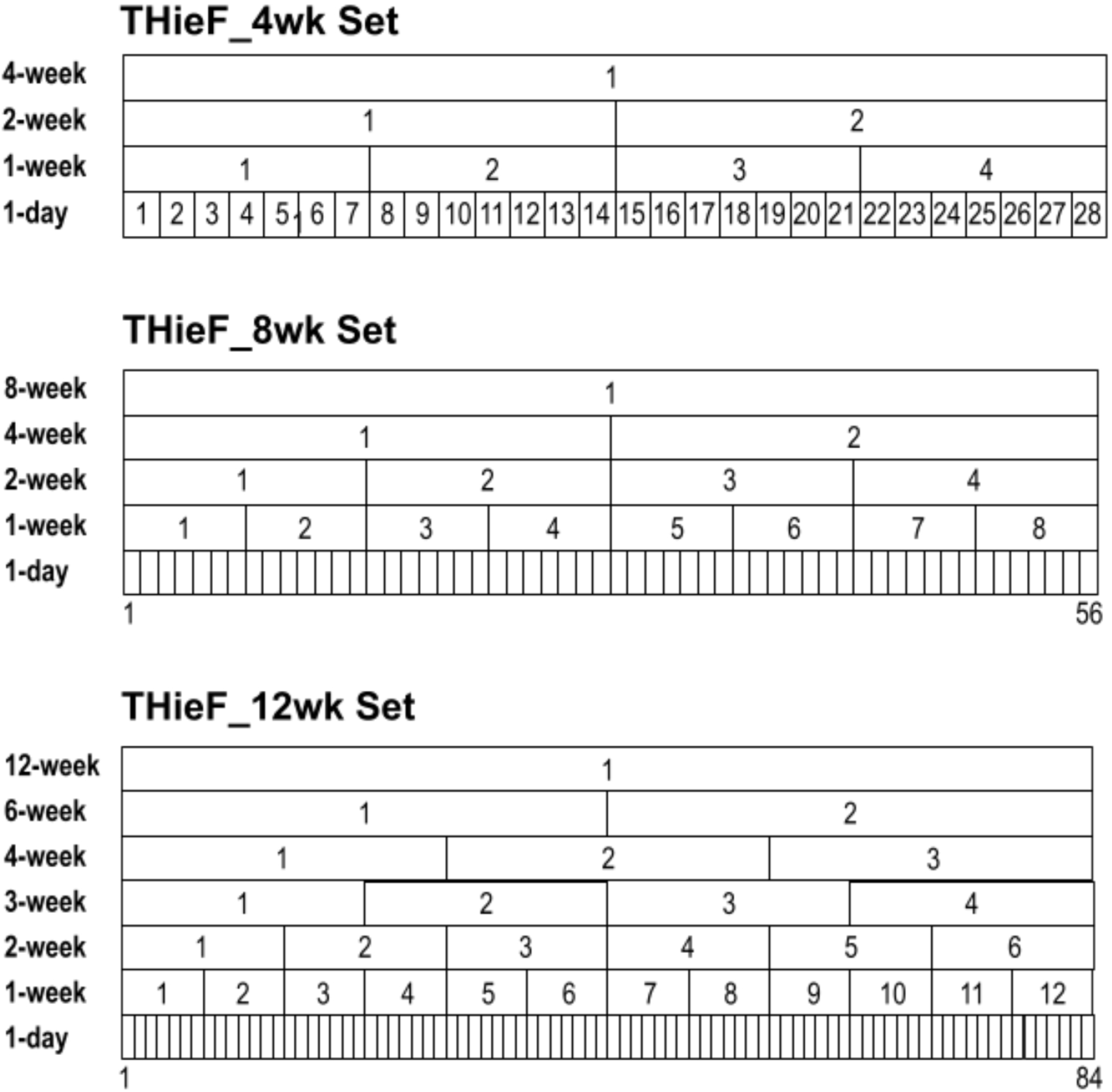
Illustration of three hierarchy schemes used to create six of the fourteen total original THieF models

Even the lowest ranking models displayed better performance compared to the COVIDhub-baseline for many locations, particularly for those with low cumulative hospitalization counts. This trend is perhaps unsurprising given that high-count settings tend to be more difficult to forecast accurately. Additionally, CU-select did not make forecasts for Puerto Rico and GT-DeepCOVID did not make forecasts for Puerto Rico or Hawaii, both of which were low-count locations that tended to be easier to predict. This may have lead these models averaged scores in the other evaluations to be slightly higher than if they had made forecasts for these locations.

## 4 Discussion

THieF is a unique forecast methodology that integrates hierarchical forecasting with forecast combination, one that showed a number of improvements in forecast accuracy over many benchmark comparison models. Such accuracy gains varied based on each model s temporal hierarchy and data transformation (or lack of thereof), with well-crafted THieF models demonstrating competitive performance. The best of these models, THieF_6wk-4root, nearly always ranked second or third in terms of WIS and MAE with typically nearer-nominal coverage rates for both scored uncertainty levels. Although it did not beat the COVIDhub-4_week_ensemble (which also ranked first when evaluating real-time COVID-19 case and death forecasting models 8), THieF_6wk-4root produced consistently accurate forecasts for incident hospitalizations using real-time truth data during the 48-week testing phase and outperformed JHUAPL-Bucky, USC-SI_kJalpha, GT-DeepCOVID, and the COVIDhub-baseline. Other well-performing models we formulated, like THieF_12wk-4root, THieF_ensemble-train3, and THieF_ensemble-mean, also produced competitive forecasts with substantial gains over the COVIDhub-baseline, GT-DeepCOVID, JHUAPL-Bucky, and USC-SI_kJalpha. The testing phase evaluations allow us to draw the following conclusions about the models we generated and the underlying methodology.

- **The THieF ensembles and original THieF models tended to be accurate compared to the baseline model**. In terms of WIS and MAE, four models30—THieF_6wk-4root, THieF_12wk-4root, THieF_ensemble-train3, and THieF_ensemble-mean-almost always beat the COVIDhub-baseline (as well as GT-DeepCOVID, JHUAPL-Bucky, USC-SI_kJalpha), while THieF_6wk-noTransform nearly always performed as well as (though sometimes better than) the baseline. This was true across evaluations split by horizon, pandemic wave, forecast date, and location; for the top three models we formulated, approximately equal improvements occurred in stable, low incidence settings and rapidly-changing, high-incidence settings. These models also demonstrated good 50% coverage and reasonable 95% coverage rates. Hence, these four models are better alternatives to the COVIDHub-baseline for predicting incident hospitalizations.
- **THieF_6wk-4root, followed closely by THieF_12wk-4root and the THieF_ensembles, yielded the best performance when compared to alternate models from the best subset**. THieF_6wk-4root, THieF_12wk-4root, THieF_ensemble-train3, and THieF_ensemble-mean ranked within the top seven for most testing phase evaluations, which compared fourteen models including the COVIDhub-baseline, JHUAPL-Bucky, USC-SI_kJalpha, GT-DeepCOVID, CU-select, and the COVIDhub-4_week_ensemble. In fact, except for the 1-week ahead horizon during times of rapid change or high incidence, THieF_6wk-4root ranked second or third in almost every analysis. It should be noted, however, that despite slightly lower rankings, especially during times of high incidence or rapid change, the THieF_ensembles showed greater stability and resistance to jumps in WIS, MAE, and PI coverage compared to THieF_6wk-4root. Overall, these models consistently had competitive performance for all four metrics out of the best subset models that moved on from the validation phase; therefore, depending on the decision-making needs involved, we conclude that any of these models would be suitable for informing COVID-19 policies.
- **The accuracy improvements of the original THieF models over the COVIDhub-baseline were likely a result of the THieF methodology’s temporal hierarchy and ensembling** During the analyses of both phases, arima_s1-noTransform, sarima_s7-noTransform, and sarima-untrained_ensemble often demonstrated poor performance, as did the other ARIMA and SARIMA models in the validation phase, and only rarely beat the COVIDhub-baseline. The best performance that arima_s1-noTransform the most accurate of the ARIMA/SARIMA models in the best subset) achieved occurred during static, low-count periods at short horizons, which was equivalent to that of higher-ranking models. We thus conclude that it was likely not the just the ARIMA based forecasters that lend most THieF_models their accuracy improvements but rather the union of hierarchical forecasting and forecast combination.
- **There is not a large difference between the trained and untrained THieF ensembles**. THieF_ensemble-train3 and THieF_ensemble-mean showed limited differences in performance during the testing phase analysis, regardless of location scale, horizon, pandemic wave, forecast date, or evaluation metric. THieF_ensembletrain3 was marginally better than the untrained THieF_ensemble-mean in most evaluations, the largest difference in relative WIS being 0.07 for any single horizon, horizon–wave, or location comparison between the models. Usually, though, the relative WIS of these two models was much closer. The by–forecast date plots provide further evidence of their similarity, as the two ensembles had almost identical WIS for every forecast date (see Figures 6 and 7). Since this trained ensemble had a small *θ* value, indicating a lighter weighting scheme, this result is not all too surprising.
- **The fourth root data transformation was essential to achieving the best results when using the THieF methodology**. We saw a range of performance demonstrated by the different THieF models during the validation and testing phase analyses, with THieF_6wk-4root and THieF_12wk-4root consistently obtaining some of the best WIS values. Conversely, THieF_6wk-noTransform had more middling performance and almost always was worse than THieF_6wk-4root. Although we did not pass on THieF_12wk-noTransform, this model ranked below its fourth root counterpart for the last 17 weeks^7^ of the validation phase. Other THieF models also outperformed the COVIDhub-baseline during the validation phase, including the THieF models with the second longest top aggregation level of eight weeks. Hence, it appears that satisfying the constant variance assumption of the underlying ARIMA model substantially improved accuracy gains from the THieF methodology.

Our results show that THieF is a powerful methodology that can model the real-world complexities of an infectious disease system, particularly if transformations are used. The initial base forecasts were created using a classic statistical model, a type of model that has shown good performance when forecasting for short horizons in outbreak settings 15, but the final reconciliation of these forecasts may be the driving factor behind THieF’s accuracy gains. Ensembling a large number of THieF models with a range of top aggregation levels achieved moderate accuracy improvements, but ultimately was not better than the top-performing original THieF models with a fourth root transformation. Combining the ARIMA and SARIMA models also did not yield a particularly accurate ensemble for the testing phase.

Further work might consider different evaluation metrics, alternative ways of aggregating forecast scores, implementing logarithmic scores, or using metrics that are not scale-dependent. The magnitude of WIS and MAE is dependent on the scale of forecasts, meaning that averages calculated from these metrics will be dominated by forecasts with large WIS and MAE values. For example, our averaged scores always had the largest contributions from longer horizons and high-count locations; that is, a model that forecasts low-count locations well but high-count locations poorly was penalized in the overall evaluation more harshly than that of a model that did the opposite. The same was true for short versus long horizons. We attempted to address this issue by considering a range of stratified analyses, but these evaluations were limited in scope. Future work that scores forecasts using other metrics that are not scale-dependent may better solve the problem of unequal contributions to aggregated scores.

The other main direction for future work is obtaining properly calibrated and truly coherent) reconciled forecasts either through sampling or by employing one of the existing probabilistic applications of THieF to forecast COVID-19 incident hospitalizations. While we generally did not observe great deviations from nominal-level coverage in our work, the THieF models coverage rates could be improved and increase overall model accuracy. Using a probabilistic implementation of THieF would generate a complete probabilistic distribution instead of using prediction intervals from point forecasts and reconciling the bounds as if they were point forecasts. Such a probabilistic implementation may be more useful or more accurate, especially in regards to improving coverage rates.

Alternative ensemble formulations may also be considered, such as: a median ensemble, a trained ensemble with a shifting *θ* value, ensembles of only THieF models with a fourth root data transform, or even ensembles of THieF implementations specifically tailored for point and probabilistic forecasting. Additionally, we might consider creating ARIMA models that train on data aggregated to different time scales instead of just using the original daily data. These new ARIMA models could also be ensembled in ways that more directly match each level of base forecaster for the various THieF model formulations to better investigate whether reconciliation is a driving factor behind the accuracy wgains we observed.

We demonstrated the utility and success of THieF and ensembles of THieF models when forecasting incident hospitalizations for COVID-19 through the results of this paper. THieF models made with fourth root–transformed data and the THieF_ensembles displayed clear improvements over the baseline model for both point and probabilistic forecasts, with THieF_6wk-4root ranking just below the top-performing COVIDhub-4_week_ensemble model and alternating second place with CU-select. Infectious disease forecasting continues to be important for acute disease outbreaks, epidemics, pandemics, and endemic diseases, and we hope that the THieF methodology will be used in such settings given its demonstrated accuracy improvements in this application.

## Supporting information

Supplemental Forecast Plots

## Data Availability

All relevant code is available at https://github.com/lshandross/covidTHieF and can be used to reproduce this manuscript and the supplementary materials. Required package versions are recorded in the renv.lock file.

## A Appendix: Supplemental materials

## Footnotes

1 Note, there was a slight exception to this restriction. See subsection 2.7 for more information.

2 Note that coherence is not an intrinsic property of all hierarchical forecasts.

3 This estimator also cannot be calculated for non-model based forecasts like expert judgements in which in-sample forecast errors may not be available [1], though this was not a concern for our work.

4 Recall that we use the 0.500 quantile forecast as the point forecast.

5 A trained ensemble created by the Forecast Hub that makes short-term forecasts for incident cases, incident and cumulative deaths, and incident hospitalizations. This model makes its forecasts by computing a weighted median of the ten best component forecasts (by WIS) in the 12 weeks leading up to the forecast date. For this model, *θ* is allowed to vary by forecast date [5, 18].

6 Note that scores for different models had to be re-aligned because teams made forecasts on different days, resulting in a mismatch between horizons for the same target and date. The scores for our models, which predicted on a Sunday, were for 3-to 30-days ahead, rather than 1-to 28 days ahead like those for some of the Forecast Hub benchmark models. This may have resulted in a slight disadvantage for our models, though only marginally.

7 We focus on the last 17 week evaluation rather than the overall one for the validation phase because the latter analysis included an anomaly when THieF_12wk-4root had an unusually large upper limit for many of its interval forecasts, which hid its otherwise better performance.

