## Supplemental Forecast Plots for "Forecasting COVID-19 with Temporal Hierarchies and Ensemble Methods"

Additional plots of 1- to 28-day ahead forecasts for daily incident COVID-19 hospitalizations for all fourteen models compared during the testing phase are provided in this supplemental document. Only five locations are shown: the entire U.S., the two locations with the highest cumulative hospitalization counts during the period of analysis (Texas and Florida), and the two locations with the lowest cumulative hospitalizations during the period of analysis (Alaska and Vermont). Note that we only plot forecasts made every four weeks, starting on the week of November 1, 2021 (the first day of the testing phase) for readability. This means that only 25% of all forecasts made and evaluated during the testing phase are shown. Forecasts are represented by a (median) point forecast, 50% and 90% prediction intervals.

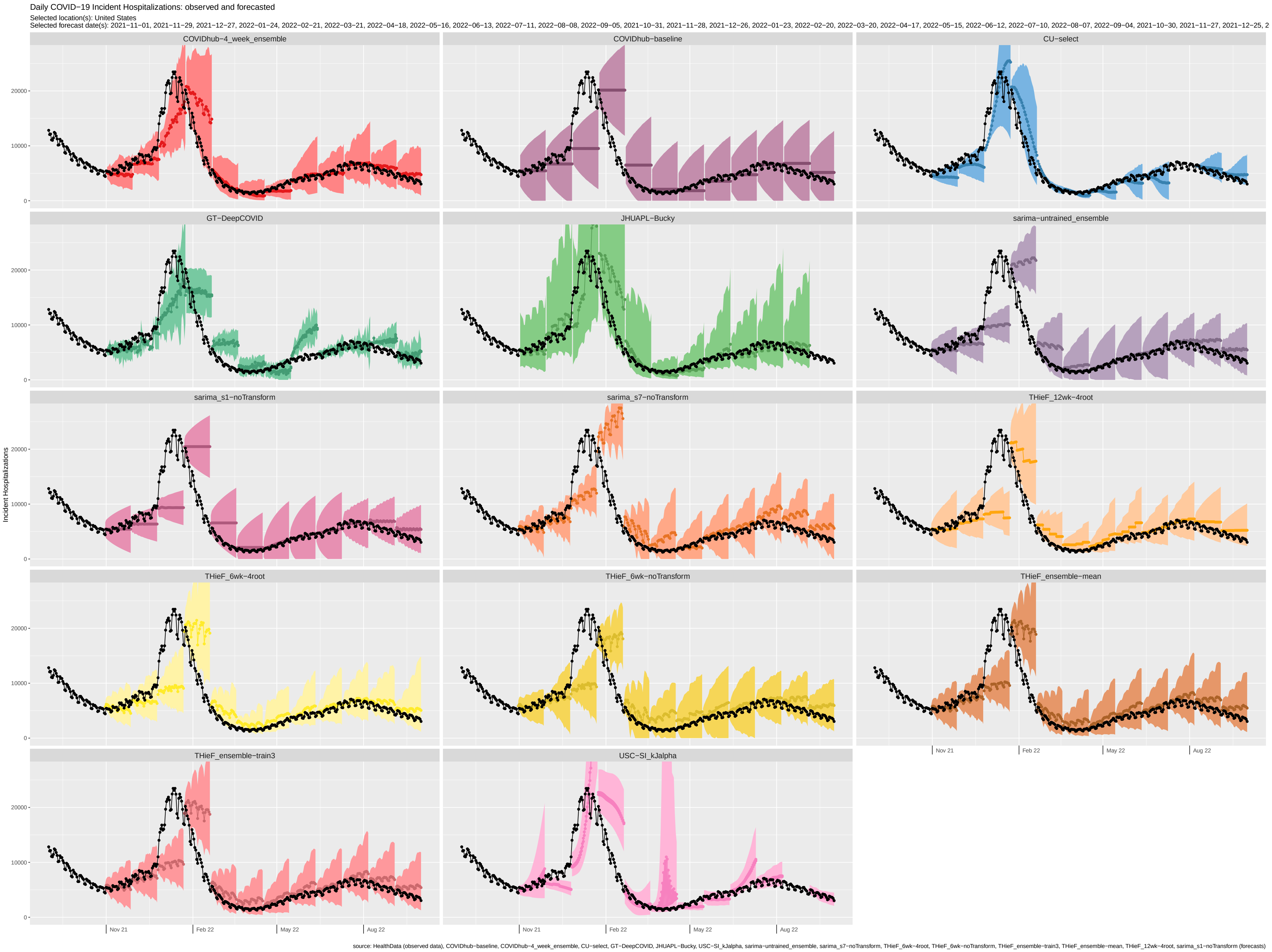
Daily COVID−19 Incident Hospitalizations: observed and forecasted
Selected location(s): United States
Selected forecast date(s): 2021−11−01, 2021−11−29, 2021−12−27, 2022−01−24, 2022−02−21, 2022−03−21, 2022−04−18, 2022−05−16, 2022−06−13, 2022−07−11, 2022−08−08, 2022−09−05, 2021−10−31, 2021−11−28, 2021−12−26, 2022−01−23, 2022−02−20, 2022−03−20, 2022−04−17, 2022−05−15, 2022−06−12, 2022−07−10, 2022−08−07, 2022−09−04, 2021−10−30, 2021−11−27, 2021−12−25, 2
COVIDhub−4_week_ensemble
COVIDhub−baseline
CU−select
GT−DeepCOVID
JHUAPL−Bucky
sarima−untrained_ensemble
sarima_s1−noTransform
sarima_s7−noTransform
THieF_12wk−4root
THieF_6wk−4root
THieF_6wk−noTransform
THieF_ensemble−mean
THieF_ensemble−train3
USC−SI_kJalpha
Incident Hospitalizations
0
10000
20000
Nov 21
Feb 22
May 22
Aug 22
source: HealthData (observed data), COVIDhub−baseline, COVIDhub−4_week_ensemble, CU−select, GT−DeepCOVID, JHUAPL−Bucky, USC−SI_kJalpha, sarima−untrained_ensemble, sarima_s7−noTransform, THieF_6wk−4root, THieF_6wk−noTransform, THieF_ensemble−train3, THieF_ensemble−mean, THieF_12wk−4root, sarima_s1−noTransform (forecasts)

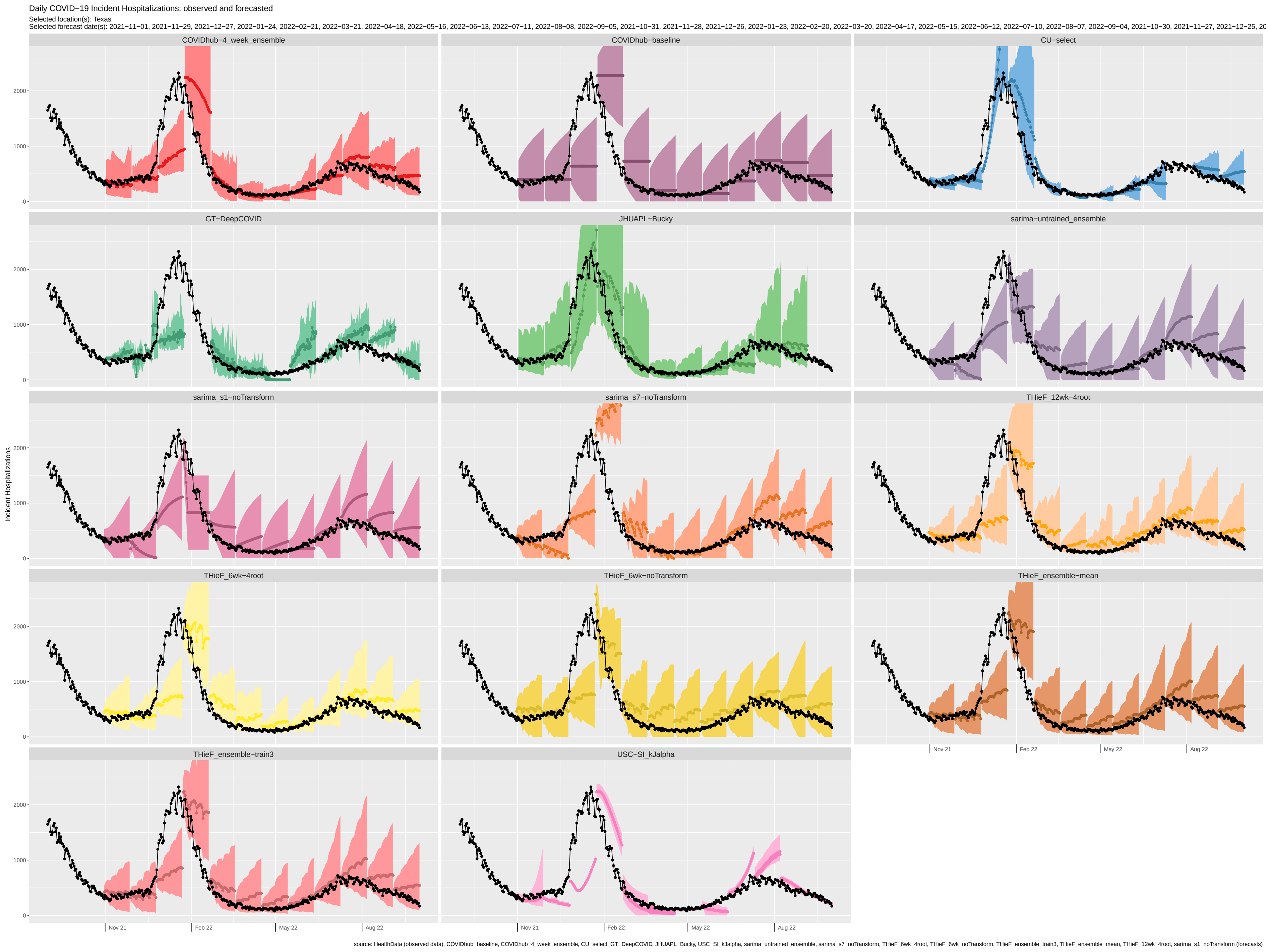
Daily COVID−19 Incident Hospitalizations: observed and forecasted
Selected location(s): Texas
Selected forecast date(s): 2021−11−01, 2021−11−29, 2021−12−27, 2022−01−24, 2022−02−21, 2022−03−21, 2022−04−18, 2022−05−16, 2022−06−13, 2022−07−11, 2022−08−08, 2022−09−05, 2021−10−31, 2021−11−28, 2021−12−26, 2022−01−23, 2022−02−20, 2022−03−20, 2022−04−17, 2022−05−15, 2022−06−12, 2022−07−10, 2022−08−07, 2022−09−04, 2021−10−30, 2021−11−27, 2021−12−25, 20
COVIDhub−4_week_ensemble
COVIDhub−baseline
CU−select
GT−DeepCOVID
JHUAPL−Bucky
sarima−untrained_ensemble
sarima_s1−noTransform
sarima_s7−noTransform
THieF_12wk−4root
THieF_6wk−4root
THieF_6wk−noTransform
THieF_ensemble−mean
THieF_ensemble−train3
USC−SI_kJalpha
Incident Hospitalizations
0
1000
2000
Nov 21
Feb 22
May 22
Aug 22
source: HealthData (observed data), COVIDhub−baseline, COVIDhub−4_week_ensemble, CU−select, GT−DeepCOVID, JHUAPL−Bucky, USC−SI_kJalpha, sarima−untrained_ensemble, sarima_s7−noTransform, THieF_6wk−4root, THieF_6wk−noTransform, THieF_ensemble−train3, THieF_ensemble−mean, THieF_12wk−4root, sarima_s1−noTransform (forecasts)

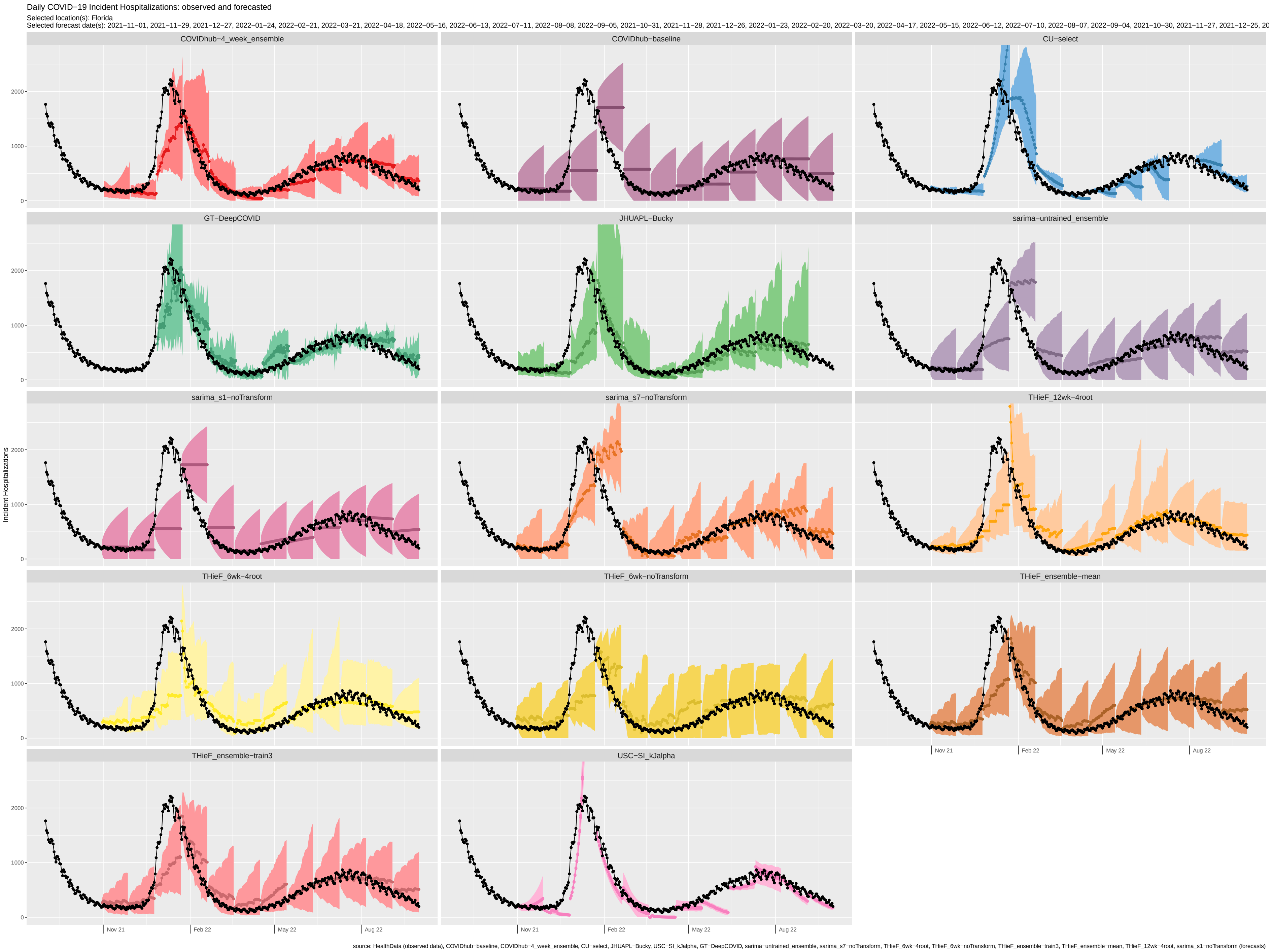

Daily COVID−19 Incident Hospitalizations: observed and forecasted
Selected location(s): Florida
Selected forecast date(s): 2021−11−01, 2021−11−29, 2021−12−27, 2022−01−24, 2022−02−21, 2022−03−21, 2022−04−18, 2022−05−16, 2022−06−13, 2022−07−11, 2022−08−08, 2022−09−05, 2021−10−31, 2021−11−28, 2021−12−26, 2022−01−23, 2022−02−20, 2022−03−20, 2022−04−17, 2022−05−15, 2022−06−12, 2022−07−10, 2022−08−07, 2022−09−04, 2021−10−30, 2021−11−27, 2021−12−25, 20
COVIDhub−4_week_ensemble
COVIDhub−baseline
CU−select
GT−DeepCOVID
JHUAPL−Bucky
sarima−untrained_ensemble
sarima_s1−noTransform
sarima_s7−noTransform
THieF_12wk−4root
THieF_6wk−4root
THieF_6wk−noTransform
THieF_ensemble−mean
THieF_ensemble−train3
USC−SI_kJalpha
Incident Hospitalizations
0
1000
2000
Nov 21
Feb 22
May 22
Aug 22
source: HealthData (observed data), COVIDhub−baseline, COVIDhub−4_week_ensemble, CU−select, JHUAPL−Bucky, USC−SI_kJalpha, GT−DeepCOVID, sarima−untrained_ensemble, sarima_s7−noTransform, THieF_6wk−4root, THieF_6wk−noTransform, THieF_ensemble−train3, THieF_ensemble−mean, THieF_12wk−4root, sarima_s1−noTransform (forecasts)

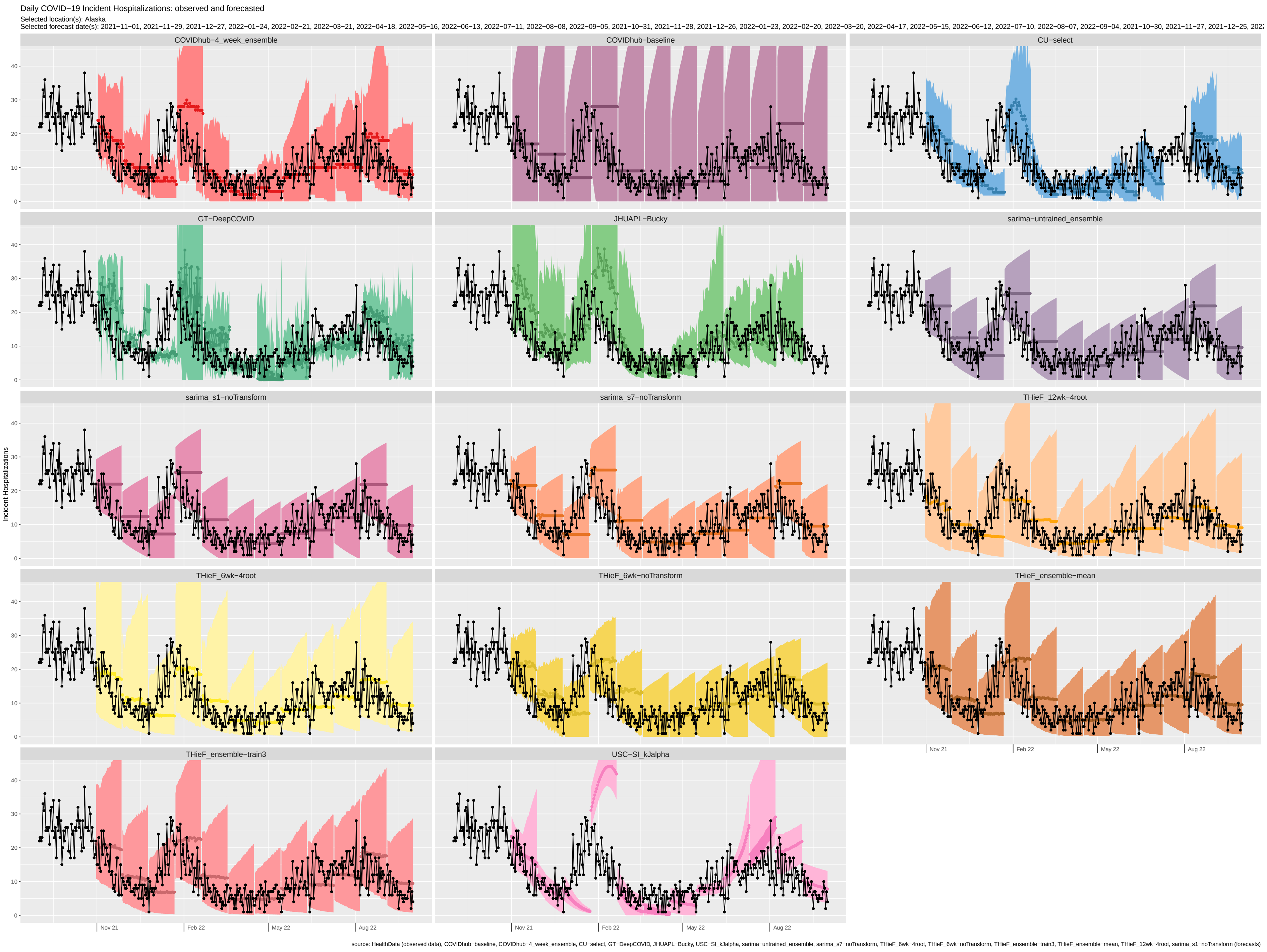
Daily COVID−19 Incident Hospitalizations: observed and forecasted
Selected location(s): Alaska
Selected forecast date(s): 2021−11−01, 2021−11−29, 2021−12−27, 2022−01−24, 2022−02−21, 2022−03−21, 2022−04−18, 2022−05−16, 2022−06−13, 2022−07−11, 2022−08−08, 2022−09−05, 2021−10−31, 2021−11−28, 2021−12−26, 2022−01−23, 2022−02−20, 2022−03−20, 2022−04−17, 2022−05−15, 2022−06−12, 2022−07−10, 2022−08−07, 2022−09−04, 2021−10−30, 2021−11−27, 2021−12−25, 202
COVIDhub−4_week_ensemble
COVIDhub−baseline
CU−select
GT−DeepCOVID
JHUAPL−Bucky
sarima−untrained_ensemble
sarima_s1−noTransform
sarima_s7−noTransform
THieF_12wk−4root
THieF_6wk−4root
THieF_6wk−noTransform
THieF_ensemble−mean
THieF_ensemble−train3
USC−SI_kJalpha
Incident Hospitalizations
0
10
20
30
40
Nov 21
Feb 22
May 22
Aug 22
source: HealthData (observed data), COVIDhub−baseline, COVIDhub−4_week_ensemble, CU−select, GT−DeepCOVID, JHUAPL−Bucky, USC−SI_kJalpha, sarima−untrained_ensemble, sarima_s7−noTransform, THieF_6wk−4root, THieF_6wk−noTransform, THieF_ensemble−train3, THieF_ensemble−mean, THieF_12wk−4root, sarima_s1−noTransform (forecasts)

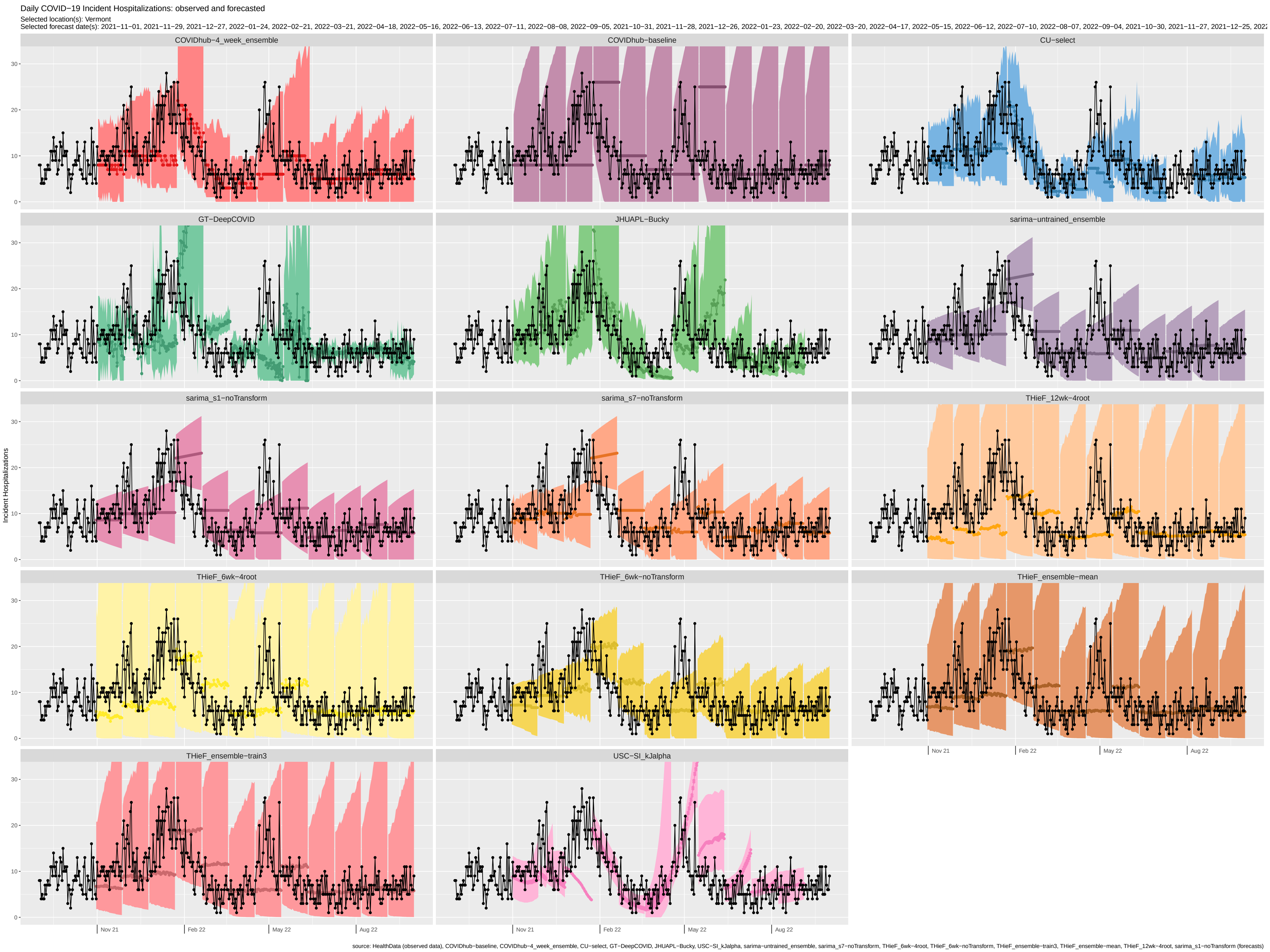
Daily COVID−19 Incident Hospitalizations: observed and forecasted
Selected location(s): Vermont
Selected forecast date(s): 2021−11−01, 2021−11−29, 2021−12−27, 2022−01−24, 2022−02−21, 2022−03−21, 2022−04−18, 2022−05−16, 2022−06−13, 2022−07−11, 2022−08−08, 2022−09−05, 2021−10−31, 2021−11−28, 2021−12−26, 2022−01−23, 2022−02−20, 2022−03−20, 2022−04−17, 2022−05−15, 2022−06−12, 2022−07−10, 2022−08−07, 2022−09−04, 2021−10−30, 2021−11−27, 2021−12−25, 2022
COVIDhub−4_week_ensemble
COVIDhub−baseline
CU−select
GT−DeepCOVID
JHUAPL−Bucky
sarima−untrained_ensemble
sarima_s1−noTransform
sarima_s7−noTransform
THieF_12wk−4root
THieF_6wk−4root
THieF_6wk−noTransform
THieF_ensemble−mean
THieF_ensemble−train3
USC−SI_kJalpha
Incident Hospitalizations
0
10
20
30
Nov 21
Feb 22
May 22
Aug 22
source: HealthData (observed data), COVIDhub−baseline, COVIDhub−4_week_ensemble, CU−select, GT−DeepCOVID, JHUAPL−Bucky, USC−SI_kJalpha, sarima−untrained_ensemble, sarima_s7−noTransform, THieF_6wk−4root, THieF_6wk−noTransform, THieF_ensemble−train3, THieF_ensemble−mean, THieF_12wk−4root, sarima_s1−noTransform (forecasts)
